# Dysregulated FOXO1 activity drives skeletal muscle intrinsic dysfunction in amyotrophic lateral sclerosis

**DOI:** 10.1101/2024.07.17.24309945

**Authors:** Mónica Zufiría, Oihane Pikatza-Menoio, Maddi Garciandia-Arcelus, Xabier Bengoetxea, Andrés Jiménez, Amaia Elicegui, Maria Levchuk, Olatz Arnold-García, Pablo Iruzubieta, Jon Ondaro, Uxoa Fernández-Pelayo, Mikel Muñoz-Oreja, Ana Aiastui, José Manuel García-Verdugo, Vicente Herranz-Pérez, Miren Zulaica, Juan José Poza, Rebeca Ruiz-Onandi, Roberto Fernández-Torrón, Juan Bautista Espinal, Mario Bonilla, Ana Lersundi, Gorka Fernández-Eulate, Javier Riancho, Ainara Vallejo-Illarramendi, Ian James Holt, Amets Sáenz, Edoardo Malfatti, Stéphanie Duguez, Lorea Blázquez, Adolfo López de Munain, Gorka Gerenu, Francisco Gil-Bea, Sonia Alonso-Martín

## Abstract

Amyotrophic Lateral Sclerosis (ALS) is a multisystemic neurodegenerative disorder, with accumulating evidence indicating metabolic disruptions in the skeletal muscle preceding disease symptoms, rather than them manifesting as a secondary consequence of motor neuron (MN) degeneration. Hence, energy homeostasis is deeply implicated in the complex physiopathology of ALS and skeletal muscle has emerged as a key therapeutic target. Here, we describe intrinsic abnormalities in ALS skeletal muscle, both in patient-derived muscle cells and in muscle cell lines with genetic knockdown of genes related to familial ALS, such as *TARDBP* (TDP-43) and *FUS*. We found a functional impairment of myogenesis that parallels defects of glucose oxidation in ALS muscle cells. We identified FOXO1 transcription factor as a key mediator of these metabolic and functional features in ALS muscle, via gene expression profiling and biochemical surveys in TDP-43 and FUS silenced muscle progenitors. Strikingly, inhibition of FOXO1 mitigated the impaired myogenesis in both the genetically modified and the primary ALS myoblasts. In addition, specific *in vivo* conditional knockdown of TDP-43 or FUS orthologs (*TBPH* or *caz*) in *Drosophila* muscle precursor cells resulted in decreased innervation and profound dysfunction of motor nerve terminals and neuromuscular synapses, accompanied by motor abnormalities and reduced lifespan. Remarkably, these phenotypes were partially corrected by *foxo* inhibition, bolstering the potential pharmacological management of muscle intrinsic abnormalities associated with ALS. The findings demonstrate an intrinsic muscle dysfunction in ALS, which can be modulated by targeting FOXO factors, paving the way for novel therapeutic approaches that focus on the skeletal muscle as complementary target tissue.

## INTRODUCTION

Amyotrophic lateral sclerosis (ALS) is a devastating disorder characterised by a progressive degeneration of upper and lower motor neurons (MN) that leads to the death of the patient within an average of 36 months from onset [10, 78]. Major symptoms, including weakness, motor disability, paralysis, and ultimately death, arise from the progressive waste of the skeletal muscle. Although MN loss and the subsequent dismantling of the neuromuscular junctions (NMJ) are considered the primary cause of muscle atrophy, recent evidence suggests that muscle pathology may be an active player in ALS disease. These data include pathological accumulation of phospho-TDP-43 within muscle fibres [42], as well as in distal axons within intramuscular nerve bundles of patients [28]. Indeed, studies carried out in SOD1 mouse models have evidenced that early metabolic alterations occur in the skeletal muscle before the onset of MN degeneration [12, 13, 33, 51]. Moreover, the overexpression of mutated SOD1 restricted exclusively to skeletal muscle cells is able to induce some of the ALS phenotypes, including loss of spinal MNs [37, 84]. Collectively, these findings support a central role of the skeletal muscle in ALS pathogenesis. This aligns with the disease-modifying effect that vigorous physical exercise appears to exert on ALS, as described in genetic and epidemiologic studies [18, 24].

Muscle fibre homeostasis in adulthood is fundamentally supported by intrinsic repair mechanisms that involve a sequence of metabolically fine-tuned steps, ranging from the activation and proliferation of skeletal muscle resident stem cells (known as satellite cells) to the differentiation and fusion into multinucleated syncytia, the myotubes, which will mature into new myofibers [59]. Several works have shown that satellite cells isolated from ALS patients exhibit changes in different myogenic markers [23, 57, 69], as well as a reduced ability to form mature myotubes under *in vitro* conditions [57, 69]. Likewise, myogenic defects have also been reported in cultured myoblasts derived from mouse models expressing mutations in either SOD1 [36, 80] or VAPB [80]. Although these myogenic defects may be legitimately considered secondary to changes in the denervated stem cell niche, the recent discovery of the moonlighting activity of *TARDBP* (a major ALS-causative gene) in muscle regeneration has bolstered the argument for the implication of myogenesis in the pathophysiology of ALS [81]. In that study, the protein expressed by *TARDBP* gene, TDP-43, was proven to be essential for skeletal muscle formation and regeneration by recruiting specific mRNAs that encode sarcomeric proteins into cytoplasmic amyloid-like granules [81]. In this line, Le Gall and collaborators demonstrated a neurotoxic paracrine role of muscle cells via secretion of extracellular vesicles in recipient cells such as the MNs, with a FUS-dependent mechanism [17]. Furthermore, two French families carrying a missense (G376V) and a C□terminal frameshift variant of TDP□43 have lately been identified to be affected by autosomal dominant myopathies [14, 90].

In light of this evidence, we postulate that abnormalities of muscle turnover in ALS might stem from intrinsic metabolic defects in muscle progenitors that eventually aggravate muscle wasting in ALS.

Based on the growing hypothesis that muscle defects contribute to MN degeneration through a dying-back mechanism [64], we sought to decipher the mechanisms underlying myogenic derangements in ALS. Our goal was to identify therapeutic targets that can mitigate or alleviate neuronal degeneration. For this purpose, we studied the contribution of two major ALS genes, *TARDBP* and *FUS*, in the metabolic regulation during myogenesis and differentiation, by using human immortalised myoblasts with induced loss-of-function (LoF) of such genes and ALS patient-derived primary myoblasts. Moreover, we analysed the functional *in vivo* consequences in *Drosophila melanogaster* ALS models with muscle-specific LoF of *TBPH* and *caz*, human *TARDBP* and *FUS* ortholog genes, respectively. Indeed, we found a general reduction in TDP-43 and FUS proteins which associated with impaired differentiation and increased expression of atrophic markers in patient-derived muscle cells, as well as repressed glycolysis as a consistent metabolic feature linking energy defects with the impaired myogenic capacity induced by TDP-

43 or FUS LoF. In addition, gene expression surveys and rescue experiments identified dysregulation of FOXO1 transcription factor as a mediator of the myogenic defects caused by TDP-43 or FUS knockdown. Inhibition of FOXO1 not only mitigated myogenic and metabolic abnormalities in ALS myoblast cells but also in other two MN disorders with muscle involvement, such as Spinal Muscular Atrophy (SMA) or Spinal Bulbar Muscular Atrophy (SBMA). Furthermore, it also rescued ALS-associated phenotypes in *Drosophila* models that bear the conditional knockdown of *TBPH* and *caz* genes specifically in muscle progenitor cells. Strikingly, muscle-nerve connections were disrupted in these fly models but corrected at the synapse level by *foxo* inhibition. Therefore, in this study, we propose FOXO transcription factors as pharmacological targets for the treatment of ALS through the mitigation of skeletal muscle metabolic and functional defects, which could improve the associated MN function.

## MATERIAL AND METHODS

### Patients, clinical management and muscle biopsies

Neurologists from the ALS Unit of Donostia University Hospital were in charge of performing diagnoses, patients’ examination and muscle biopsies. Samples and clinical data from patients included in this study were provided by the Basque Biobank (www.biobancovasco.org) and processed following standard procedures with appropriate approval by the Clinical Research Ethics Committees of the Basque Country and Donostia University Hospital (codes: ALM-EMP-2015-01, PI2016075, PI2019198). Informed consent was obtained from all the subjects. This research project was conducted following the requirements of The European Regulation of Data Protection and Digital Rights guarantee. All samples were anonymized, only keeping the necessary gender and age information.

All muscle biopsies were performed using the open minor surgery technique at the neurologist’s consulting room, except for the *post mortem*, which was recovered at the autopsy room. 21 muscle biopsies were used in this study, as summarised next (Table 1): 8 Controls (CTRL), 30-75 years-old; 3 Familial ALS (fALS), 45-71 years-old; 5 Sporadic ALS (sALS), 47-66 years-old; 3 MN disorders (SMA3, caused by mutations in SMN1; SBMA, also known as Kennedy’s disease, related to pathogenic GCC expansions in the androgen receptor; CMT4, Charcot-Marie-Tooth type 4, a recessive and demyelinating disease); and 2 muscular disorders with protein aggregates (sporadic Inclusion Body Myositis-IBM and a vacuolar myopathy related to Valosin-Containing Protein-VCP). CMT4 myoblasts were provided by the MYOBANK-AFM (Myology Institute) (Table 1).

**Table 1.**
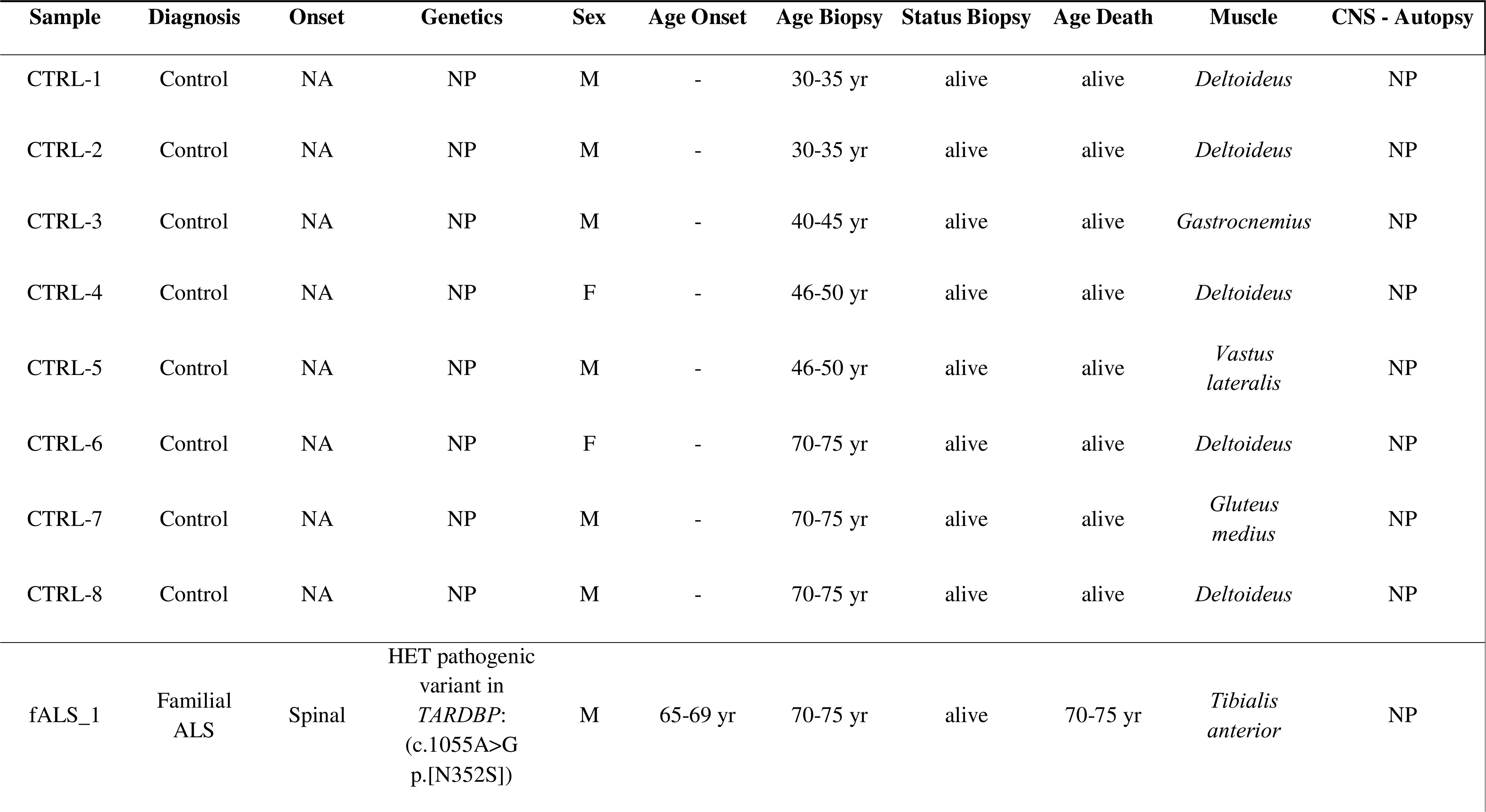
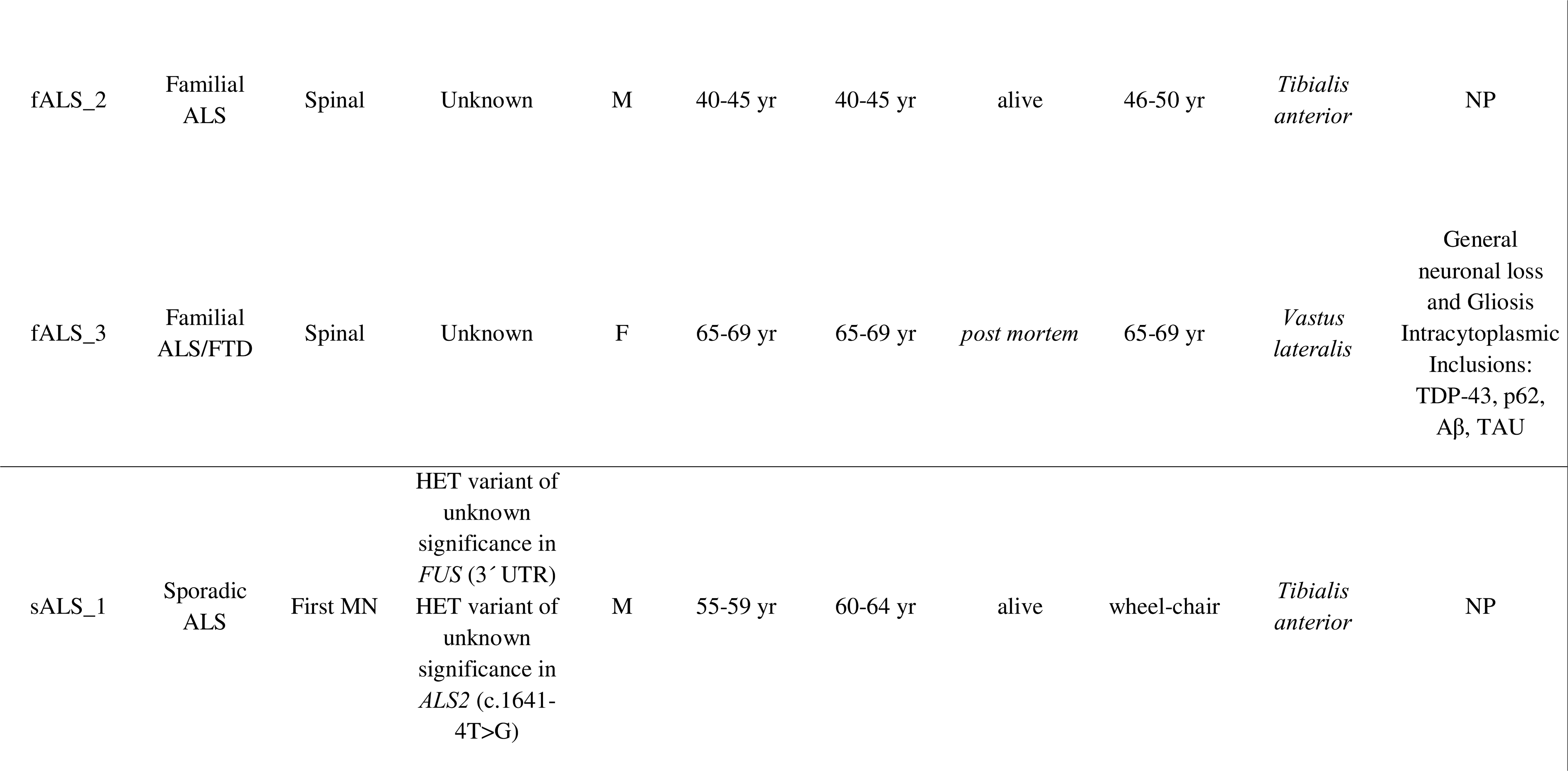
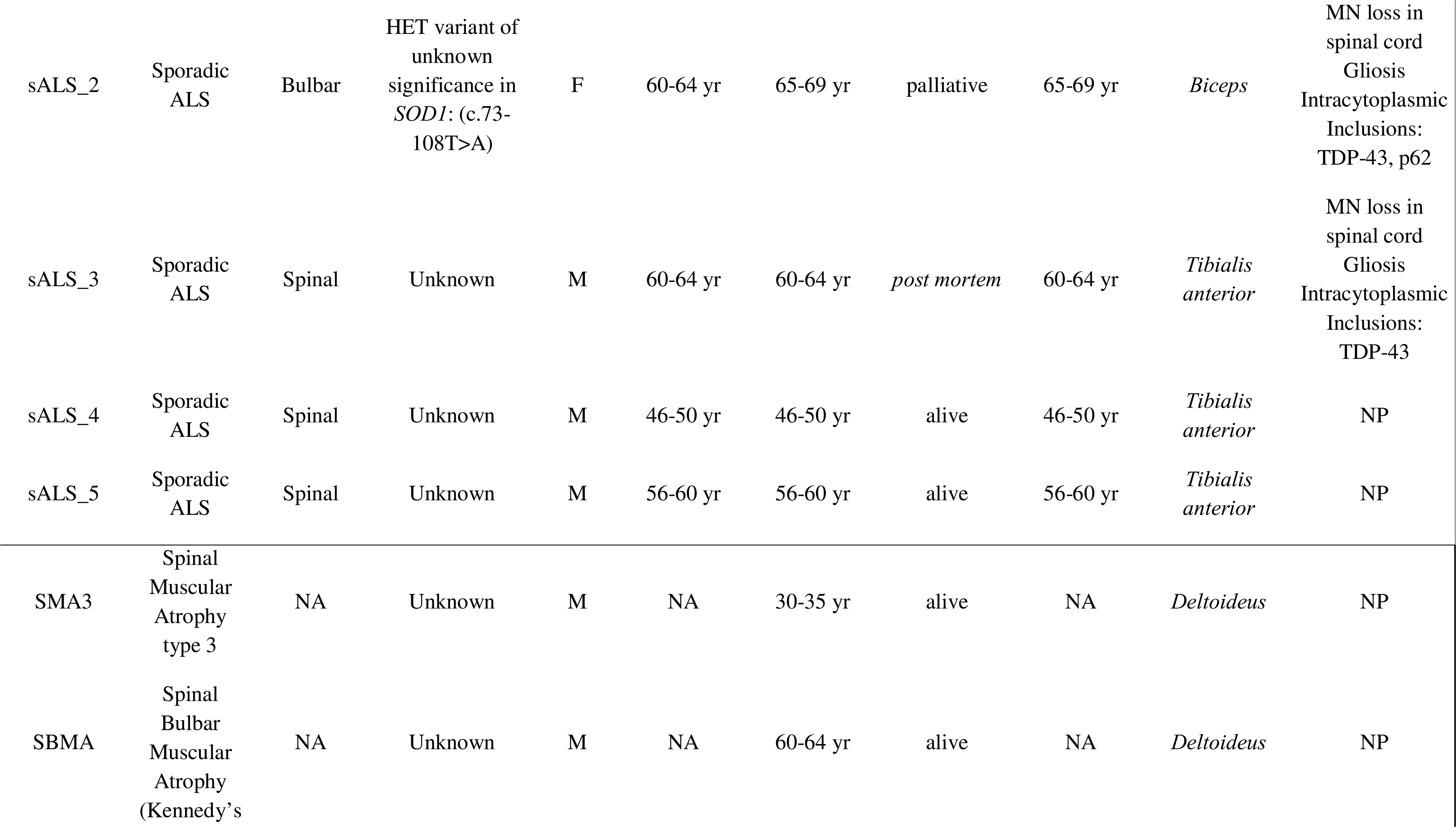
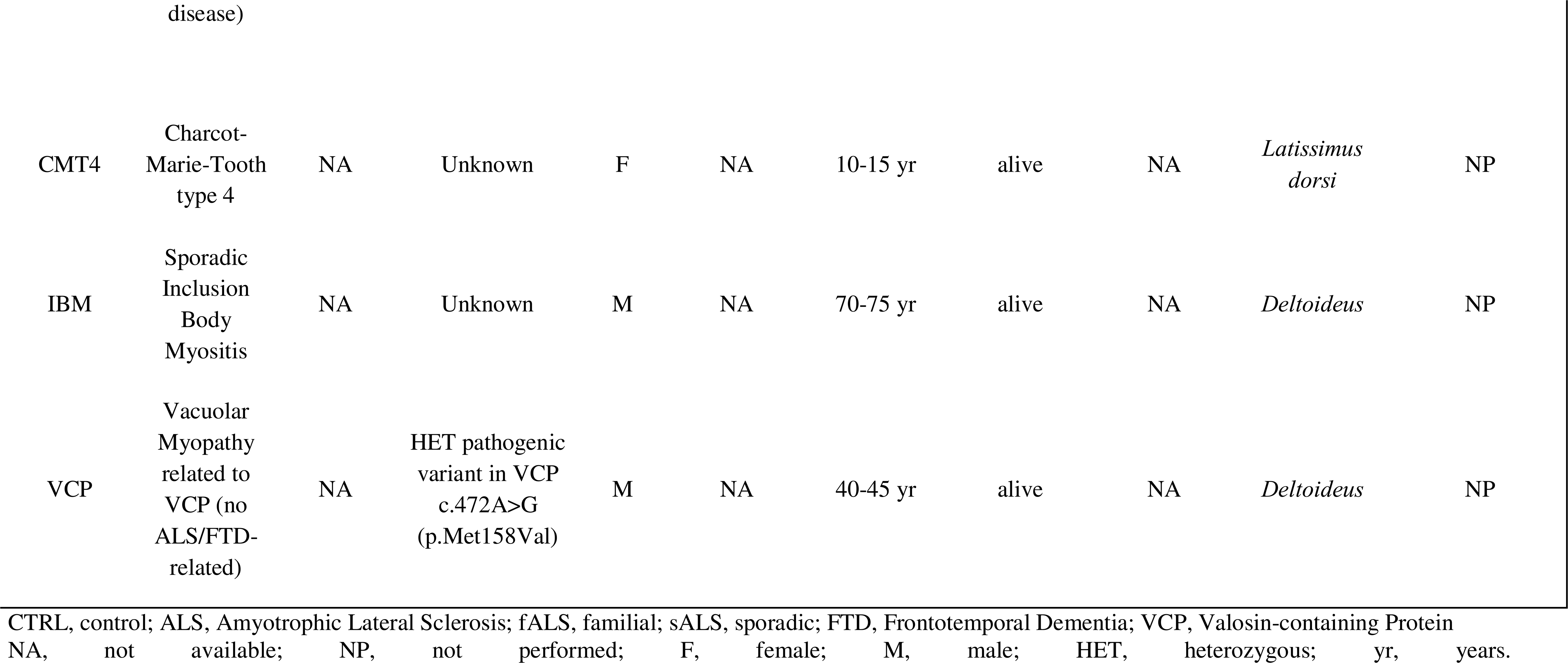
Clinical characteristics of participants and their biological samples.

### Haematoxylin and Eosin Staining in histological cryosections

Muscle biopsies were recovered and cleared of vessels and connective tissue. Several pieces from each biopsy were snap-frozen in liquid nitrogen-cooled isopentane to be processed for histological analysis. 8 µm cryosections were obtained and fixed in 4% paraformaldehyde (PFA) and processed for Haematoxylin and Eosin staining as previously described [65]. The histological preparations were mounted with DPX Mountant (Sigma-Aldrich) and scanned at 10X magnification on Pannoramic MIDI II (3DHistech).

### Cell culture assays

#### Primary myoblasts from patients’ muscle biopsies

Human proximal muscle biopsies were minced and explants were cultured to obtain myogenic precursors in a monolayer as described previously [61]. To obtain highly purified myoblasts (CD56+), primary cultures were sorted by immunomagnetic selection based on the presence of the early cell surface marker CD56 (separator and reagents from Miltenyi Biotec, Bergisch Gladbach, Germany). Cellś myogenicity was confirmed by Desmin immunolabeling, and all cultures were used at *passages* 3 and 4. Finally, myoblasts were grown up to the desired density in an incubator at 37°C, with 5% CO_2_ and a humidified atmosphere. The proliferation medium used from this moment was the Skeletal Growth Medium (SGM, PromoCell), supplemented as stated below. For *post mortem* samples, different reports have demonstrated that muscle progenitor cells, while retaining their regenerative capacity, grow and differentiate as well as those obtained from fresh biopsies [30, 50, 54].

#### Immortalised human control myoblasts (line 8220)

The immortalised human myoblast control cell line 8220 was generated at the Platform for the Immortalization of Human Cells / Myology Institute (Paris, France) and kindly provided by Dr. Vincent Mouly [29]. These myoblasts were grown and transduced as previously described [29], with minor modifications. Briefly, the proliferation medium consisted of SGM supplemented with 10% FBS, 1% Glutamax and 1% gentamicin (all from Gibco-Invitrogen). Gene silencing was carried out by transduction with lentiviral particles containing short hairpin RNA (shRNA). The plasmids used to create these particles were: SHC001 (shRNA Empty Vector Control Plasmid DNA), TRCN000016038 (human, *TARDBP* MISSION shRNA), TRCN0000288639 (human, *FUS* MISSION shRNA), TRCN0000010333 (human, *FOXO* MISSION shRNA) (MISSION® pLKO.1-Pure) (all from Sigma-Aldrich). Viral particles were produced and titled by the Viral Vector Unit of CNIC (Madrid, Spain). Myoblasts were seeded in 6-well plates at a density of 80,000 cells/well, and the next day viral particles were added at a MOI of 10. 24 hours after transduction, the medium was removed and replaced by fresh SGM proliferation medium. To verify the degree of gene silencing, TDP-43 and FUS protein expression was measured by Western blot.

#### Induction and assessment of myogenic differentiation

Either primary or immortalised myoblasts were cultured in SGM medium supplemented with 10% FBS, 1% Glutamax and 1% gentamicin, and differentiation occurred spontaneously without medium refreshing by cell-cell contact when cultures were at high confluence. The differentiation index was calculated as the percentage of nuclei in MyHC-positive cells out of the total (DAPI-positive) nuclei. The fusion index was calculated as the percentage of nuclei fused into myotubes (MyHC-positive syncytia with at least two nuclei) out of the total nuclei. Myogenicity was assessed as the percentage of MYOD-positive or MYOG-positive nuclei, out of the total nuclei. Over 3 wells and 3-10 fields were evaluated per each group.

#### 2-Deoxy-D-glucose (2-DG) treatment

For Western blot analysis, cells were seeded at 60% of confluence in 6-well plates in 2 mL of SGM, allowing them to adhere overnight (O/N). The next day, cells were treated with 100 µM 2-DG and kept for 4 days, when untreated cells were at 100% of confluence. For immunofluorescence analysis, cells were seeded at 100% of confluence in µ-slide wells (Ibidi) in 60 µl of SGM, and allowed to adhere O/N. The next day, cells were treated with 100 µM 2-DG for 48h.

#### Proliferation curves

2,000 cells/well of primary human myoblasts or 2,500 cells/well per silenced human control myoblasts (shRNA, shTDP-43 and shFUS) were plated in µ-slide wells (Ibidi), and fixed at three time points: day 1, day 3 and day 5. DAPI staining was used to quantify cell numbers at all times. Images were taken with ZEISS LSM 900 confocal microscope, and quantification of DAPI+ cells was performed with ImageJ v2.9.

#### iFOXO1 treatment

33 nM (IC50) of FOXO1 inhibitor *AS1842856* (Astella Pharma; Shelleck; # S8222) was used in all *in vitro* experiments. For Seahorse experiments, myoblasts were treated with iFOXO1 inhibitor after seeding them in a 96-well XF cell culture microplate (Agilent) for 24 h. For differentiation experiments, after fusion, silenced or ALS patient-derived myoblasts were kept with the inhibitor for 8 days. For proliferation experiments, cells were treated with iFOXO1 or the corresponding amount of DMSO, refreshing it every 3 days. Finally, for *in vivo* experiments in *Drosophila*, 30 µM iFOXO1 was diluted in the food.

### *Drosophila melanogaster* models and functional assays

One *Drosophila* strain where Cabeza (*caz*) gene is silenced in muscle cells (*UAS-Dicer2-icaz-Mef2-GAL4,* iCaz) and three *Drosophila* strains where *TBPH* gene is silenced at different locus in muscle cells were generated: *UAS-Dicer2-iTBPHattp40-Mef2-GAL4* (iTBPH^attp40^); *UAS-Dicer2-iTBPHp(GD6943)-Mef2-GAL4,* (iTBPH^p(GD6943)^); and *UAS-Dicer2-iTBPHp(pkk108354)-Mef2-GAL4*, (iTBPH^pkk108354^) were generated. As control flies, the strain *UAS-Dicer2-+-Mef2-GAL4* was used. To generate these genotypes, stocks from Vienna Stock Center (VDRC, Austria) (*caz*, #100291; *TBPH^GD6943^* #38377 and *TBPH^pkk(108354)^* #104401) and Bloomington Stock Centre (Indiana, USA) (*TBPH^attP40^* #39014 and *Dicer2-Mef2* #25756) were acquired. Flies were housed at 23°C, 70% humidity and 12 h/12 h light/darkness cycle. For longevity assays, one hundred adult flies (five per tube) were selected from each strain. Dead flies were counted every 2 days. Kaplan–Meier method was used to plot the results. A log-rank test was used to analyse results and Bonferroni correction for multiple comparisons. For locomotor activity, adult flies on days 5, 10 and 15 were analysed in groups of five. They were placed in a tube with a line drawn outside the tube at 8 cm height from the bottom. The number of flies that passed the line in 10 seconds was counted (3 trials per tube). Adult pharate survival was expressed as a percentage of the adult flies counted over the total number of pupae in each tube where adult pharate survival of control flies was around 100%. 6 tubes were counted per group in i*TBPH^pkk(108354)^* flies.

### Western blotting

Bulk protein extraction from cell cultures was done with a sample buffer containing 62 mM Tris (pH 7.5), 5% glycerol, 2% SDS, 5mg/ml bromophenol blue and 5% β-mercaptoethanol. In some experiments, cell fractionation was carried out using the NE-PER kit (Thermo Scientific, Catalog Code 78833) before adding the sample buffer. Samples were run in Criterion^TM^ gels (Bio-Rad, Life Sciences, CA, USA) by SDS-PAGE electrophoresis. Proteins were transferred to BioTrace^TM^ Nitrocellulose membranes (Pall Corporation, NY, USA). Primary antibodies are summarised in Supplementary Table 1. Secondary antibodies conjugated to Alexa fluor plus fluorophores (Invitrogen) were diluted to 1/10,000 in TBS-T (tris buffered saline with 1:1000 Tween 20) with 5% BSA (both from Sigma-Aldrich). Bands were visualised using iBright FL1000 Imaging System and quantified with Image Studio Lite v5.2. β-actin or β-tubulin were used as loading control. Results were expressed as optical density (O.D.).

### Immunofluorescence

Myoblasts or myotubes grown on µ-slide or µ-plate wells (Ibidi) were fixed after several washes in PBS, with 4% paraformaldehyde (PFA) for 10 min. After several washes, fixed cells were blocked in a solution containing 5% BSA, 2% horse serum (HS, Gibco-Invitrogen), 0.02% sodium azide and 0.5% Triton X-100 in TBS-T, for 30 min. Cells were incubated with primary antibodies O/N at 4°C diluted in blocking solution. Primary antibodies are summarised in Supplementary Table 1. The next day, cells were rinsed three times in TBS-T. Secondary antibodies conjugated to Alexa fluor plus fluorophores (Invitrogen) were diluted to 1/1,000 in blocking solution and incubated for 2 h at room temperature (RT) in the dark. DAPI was added together with the secondary antibody to visualise the cell nuclei. ZEISS LSM 900 laser scanning confocal microscope (Carl Zeiss, Inc., Germany) and Fluorescent Nikon 80i microscope were used to analyse the immunostaining. Levels of nuclear protein expression were assessed by quantifying the integrated densities of nuclei by ImageJ v2.9 using DAPI staining to delimit nuclei. For the assessment of MYOD-, MYOG- and FOXO1-positive nuclei, integrated density values above the cut-off point (set as the value of mean +1*SD of control cells) were considered positive. For the assessment of nuclear depletion of TDP-43 or FUS, nuclei with integrated density values below the cut-off point (set as the value of mean −1*SD of control cells) were considered negative. To evaluate the nuclear expression of FOXO3, the integrated density values of each nucleus were quantified and plotted.

### Transmission electron microscopy

Cells were postfixed in 1% osmium tetroxide, 7% sucrose in 0.1 M PB for 30 min at RT, washed in deionized water, and partially dehydrated in ethanol. Cells were then stained in 2% uranyl acetate in 70% ethanol in the dark for 2 h 30 min at 4°C. Cells were further dehydrated in ethanol, and infiltrated O/N in Durcupan ACM epoxy resin (Fluka, Sigma-Aldrich, St. Louis, USA). Following resin hardening, embedded cell cultures were detached from the chamber slide and glued to resin blocks. Serial semi-thin sections (1.5 µm) were cut with an Ultracut UC-7 ultramicrotome (Leica, Heidelberg, Germany) and mounted onto glass microscope slides and lightly stained with 1% toluidine blue. Selected semi-thin sections were glued with Super Glue-3, Loctite (Henkel, Düsseldorf, Germany) to resin blocks and subsequently detached from the glass-slides by repeated freezing (in liquid nitrogen) and thawing. Ultra-thin sections (70-80 nm) were obtained with the ultramicrotome from detached semi-thin sections, and further stained with lead citrate (Reynolds’ solution) [60]. Finally, cells were imaged at 80 kV on a FEI Tecnai G^2^ Spirit transmission electron microscope (FEI Europe, Eindhoven, Netherlands) equipped with a Morada CCD digital camera (Olympus Soft Image Solutions GmbH, Münster, Germany).

### Histopathological analysis of *Drosophila melanogaster* muscle

Immunofluorescence was used to study the motoneurons and the integrity of the neuromuscular junction in the indirect flight muscles of the iTBPH^pkk108354^ and control *Drosophila* models. Flies were anaesthetised with CO_2_-enriched air. Fly thoraces were isolated and immersed into 70% EtOH for 1 min. and placed in PBS for dissection. Samples were fixed with 4% PFA for 30 min. After 3 washes in PBS with 0.5% Triton X-100 (PBS-T), samples were immersed into liquid nitrogen for 10 seconds and transferred into cold PBS. Thoraces were divided longitudinally, washed 4 x 5 min in PBS-T, and incubated for 30 mins in blocking solution (5% BSA and 0.02% NaN_3_ in PBS-T) at RT. Subsequently, thoraces were incubated O/N at 4°C with the primary antibody solution (HRP and nc82 (Supplementary Table 1) in blocking solution at 4°C. After 4 x 20 min washes with PBS-T, samples were incubated O/N at 4°C with the secondary antibodies conjugated to Alexa fluor plus fluorophores (Invitrogen) and Invitrogen™ Alexa Fluor™ 647 Phalloidin 1:1000 in blocking solution. Samples were mounted in ProLong^TM^ Diamond Antifade Mountant (Invitrogen). Images were obtained in a confocal microscope LS900 (Zeiss) by z-stack scanning (total thickness of 15 μm with a step size of 0.5 μm per slice) of the whole-mount thoraces (halves). Quantification of muscle area covered by HRP staining was carried out by using ImageJ v2.9, whereas quantification of nc82-positive synaptic boutons size was performed by using SynPAnal software [11].

### RNA-seq analysis

#### RNA preparation and RNA-seq performance

Total RNA was extracted from three independent experimental replicates using the RNeasy Mini Kit (Ref. 74104, QIAGEN), followed by DNase I treatment. RNA integrity (3 *shRNA*, 3 *shTDP-43*, 3 *shFUS*) was assessed (Agilent 2100 Bioanalyzer) before ribosomal RNA depletion (Illumina Ribo-Zero Plus) and library preparation following a Stranded Full-length Total RNA procedure. Sequencing was performed on Illumina NextSeq2000 platform at a sequencing depth of at least 20M paired-end 100 nucleotide long reads (PE100). After sequencing, raw reads were filtered to remove adaptor sequences and low-quality reads (phred quality score below Q33).

#### RNA-seq data processing and analysis

RNA-seq reads were quasi-mapped to the reference transcriptome hg38 and the abundance of each transcript was quantified using Salmon v.1.5.2. with default options for paired-end reads [53]. Transcriptomes were indexed using the decoy-aware method in Salmon. Output quants files were then processed in R Studio v.4.0.4 (RStudio Team, 2020; http://www.rstudio.com/). Tximport package was used for non-normalized gene-level counts estimation from transcript-level counts using the “*bias corrected counts without an offset*” method [73]. R package Normalization was performed using DEseq2 R package [34] to account for technical variability and estimate size factors for accurate comparisons of gene expression between samples. A log2 fold change (Log2FC) threshold of greater than or equal to 1 (corresponding to a fold change of 2 or greater) was used to identify genes with statistically significant changes in expression after adjusting for multiple testing using the Benjamini-Hochberg method (adjusted *p*-value < 0.001). Functional pathway analysis was performed by Hallmark Pathway enrichment of differential gene expression signatures using GSEA v.4.3.2 as previously described [77]. Classic weighting was used as the enrichment statistics, and the permutation type was based on a gene set, using a total of 1,000 iterations. A gene set is considered significantly enriched if its Normalized Enrichment Score (NES) has a False Discovery Rate (FDR) *p*-value below 0.05 (Benjamini-Hochberg method) and a nominal *p*-value below 0.001. A GSEA score (based on multiplying NES by −log_10_FDR *p*-value) was calculated for each gene set, and their values for each gene list were plotted.

Transcription factor (TF) activity analysis was performed using ISMARA (Integrated System for Motif Activity Response), an online tool for automatically inferring regulatory networks from gene expression data [1]. Quality and adaptor trimmed fastq.gz RNA-seq files were uploaded to www.ismara.unibas.ch for processing, followed by sample average. For each enriched TF motif identified, the ISMARA score was calculated based on the z-score and average gene target expression change.

#### Data availability

Sequencing data have been deposited in GEO under accession code GSE227573.

### Analysis of metabolic flux by SeaHorse

Metabolic pathways were studied using the Seahorse XF96 Extracellular Flux Analyzer (Agilent). This equipment allows simultaneous study of the two main energy pathways, mitochondrial respiration and glycolysis, in living cells and in real time. The analyzer measures the ratio of oxygen consumption (OCR, Oxygen Consumption Rate), as an indicator of mitochondrial respiration; and the ratio of extracellular acidification (ECAR, ExtraCellular Acidification Rate), as an indicator of glycolysis and lactate production. The day before the experiment, 15,000 primary myoblasts/well were seeded in a 96-well XF cell culture microplate (Agilent) to reach 100% confluence. Before starting with the Glyco-stress experiment, media was changed to glucose-free Seahorse XF MEM media (pH 7.4) (Agilent #103575-100) for 1 h, and it was incubated at 37°C in an incubator without CO_2_. Then, continuous measurements of ECAR or OCR started. The data were normalised to cell number, which was measured by Cresyl violet staining (0.1% solution for 15 min).

All metabolic tests using the Seahorse technique were performed when myoblasts were at a confluence of 100%. For the study of metabolism across muscle differentiation, cell seeding was carried out at different stages (myoblast, myocyte, 3-day, and 8-day myotubes) in order to obtain, in the same Seahorse plate, the different phases of the process. One hour before the test, cells were incubated at 37°C without CO_2_ and in the glucose-free Seahorse XF MEM medium at pH 7.4.

### Statistical analysis

Unless otherwise indicated, data were analysed and visualised by GraphPad Prism 7. Normality was verified by the Shapiro-Wilks test (*p*<0.05). The choice of statistical test for group comparisons was guided by the normality of the data distribution. For pairwise comparisons between two groups, the Mann-Whitney *U* test was employed when the data exhibited a non-normal distribution. In contrast, when the data followed a normal distribution, Student’s t-test was utilized. For comparisons involving more than two groups, one-way or two-way ANOVA was followed by Dunnett’s test. Unless otherwise indicated, the data are expressed as the mean ± the Standard Error of the Mean (SEM).

## RESULTS

### Myogenesis is impaired in ALS patients’ cells independently of the motoneuron input

Histopathological analysis of muscle biopsies from age-matched control (CTRL) and ALS patients, familial (fALS) and sporadic (sALS) patients, revealed chronic muscle affectation, including myofiber atrophy, a few necrotic myofibers and either pyknotic or centralized nuclei and cell infiltration in ALS cases (Supplementary Fig. 1). These findings indicate an already impaired muscle homeostasis across all ALS biopsies analysed. To further investigate these alterations, we isolated muscle progenitor cells derived from 16 different subjects (8 CTRL, 3 fALS and 5 sALS cases; Table 1) and performed *in vitro* studies, devoid of inputs from MNs or any other external stimuli able to modify the intrinsic properties of muscle cells. This approach allowed for a comprehensive characterization of the myogenic process *per se* in ALS. We first assessed the myogenic capacity of the expanded cells through differentiation assays. Our results showed that most ALS myoblasts exhibited impaired differentiation, characterized by decreased differentiation and fusion indices, as well as the formation of fewer and thinner myotubes, compared to controls (Fig. 1a, b). Notably, one patient’s myoblasts (fALS_2) demonstrated elevated and comparable differentiation levels to controls. Interestingly, this patient’s disease progression was particularly slow (Table 1). It is noteworthy that myogenic differentiation capacity is variable among controls, and this variability can be attributed to age, as there was a negative correlation between differentiation index and age (Fig. 1c). However, in ALS patient myoblasts myogenesis was affected regardless of age (Fig. 1c). Furthermore, myogenesis was already impaired before differentiation and fusion events began, as evidenced by a significant and consistent reduction in the expression of MYOD (Supplementary Fig. 2a), the key transcription factor regulating myoblast differentiation, and myogenin (MYOG) (Supplementary Fig. 2b), a crucial factor for myoblast differentiation and fusion. Consistently, fALS_2 cells did not display altered MYOG expression, which may account for the lack of myogenic defects observed in these cells (Supplementary Fig. 2a).

**Figure 1.**
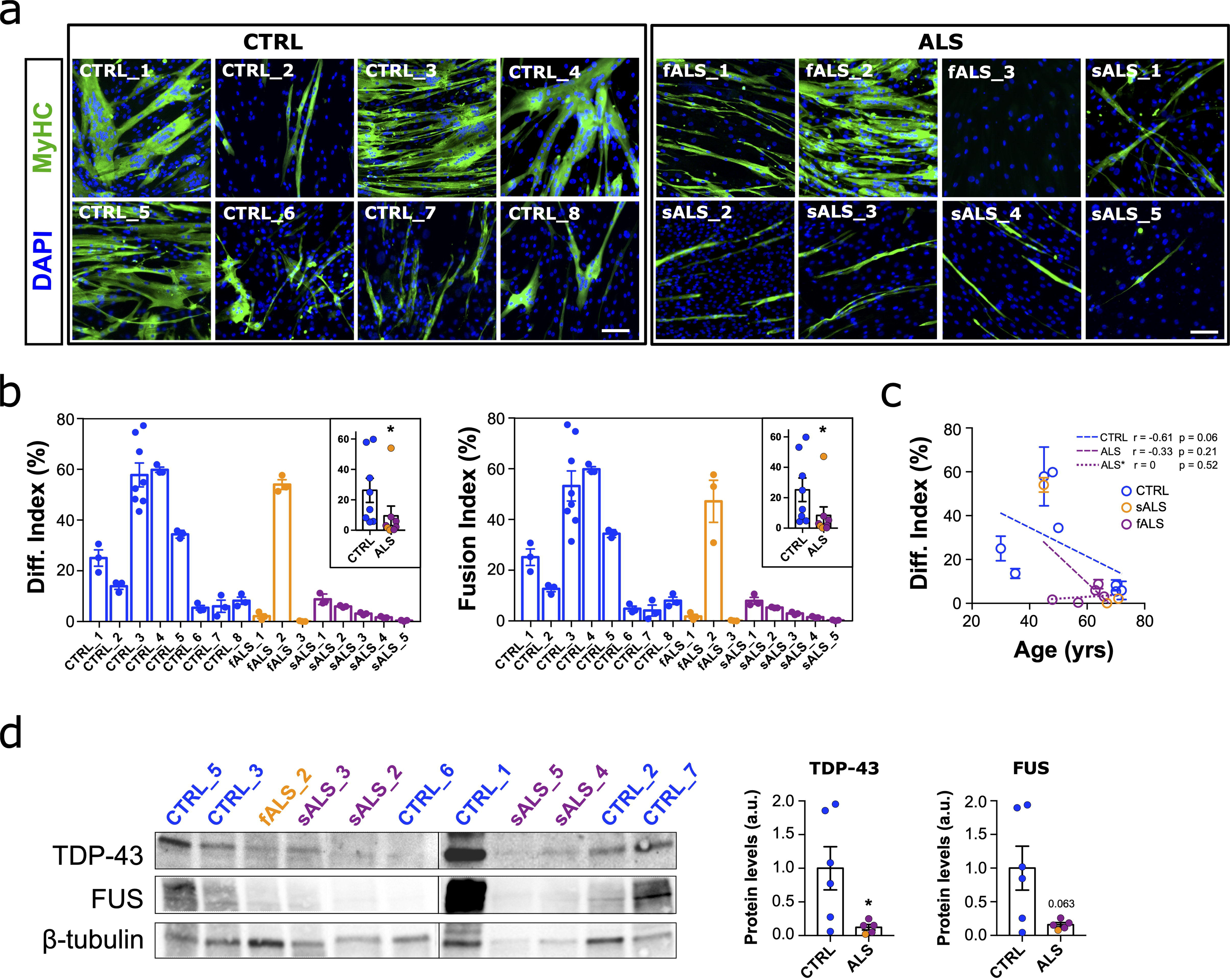
Pathological and functional abnormalities of primary myoblasts from ALS patients. **a** Representative images of primary myoblasts from ALS patients, both familial (fALS) and sporadic ALS (sALS) and healthy controls (CTRL) stained for Myosin Heavy Chain (MyHC, green) and DAPI (nuclei, blue). **b** Quantitative scatter plots with bar graphs showing fusion and differentiation indices of each individual’s cells from at least n=3 independent differentiation assays. The inner plots depict the comparison of the average values of each ALS patient group (fALS and sALS) with the average values of the CTRL group. **c** The correlation plot shows a negative association between age and the differentiation index in CTRL myoblasts (r = −0.61, p = 0.059), indicating that as age increases, the differentiation index tends to decrease. In contrast, there is no significant association in ALS myoblasts (r = −0.33, p = 0.214). When the outlier value (fALS_2), identified by Grubb’s test, is removed, the correlation in ALS* myoblasts becomes r = 0 (p = 0.520), suggesting no relationship between age and differentiation index. Bar graphs represent mean ± SEM. \**p*<0.05 compared to the CTRL group via Mann-Whitney *U* test. Scale bar: 100 μm. **d** Representative Western blot images of TDP-43 and FUS proteins in CTRL and ALS proliferative myoblasts, and scatter dot-plots with bar graphs showing the quantification of protein levels. n=3 per group. \**p*<0.05 compared to the CTRL group via Mann-Whitney *U* test.

On top of that, ALS-derived myoblasts exhibited decreased proliferation rates overall (Supplementary Fig. 2c, d). Furthermore, we analysed regulators of atrophy pathways in the skeletal muscle, such as the muscle-specific E3 ubiquitin ligases MURF1 and MAFbx/Atrogin-1, and found an increase in ALS samples (Supplementary Fig. 2e), suggesting an activation of the ubiquitin/proteasome system, leading to protein degradation.

Finally, the levels of the two proteins closely associated with ALS pathology, TDP-43 and FUS, were evaluated. We observed a significant decrease in total TDP-43 levels and decreased FUS levels by immunoblotting (Fig. 1d). Both TDP-43 and FUS are nuclear ribonucleoproteins that shuttle between the nucleus and cytoplasm. Under certain pathological conditions or in response to stress, they can accumulate in the cytoplasm [7]. However, in myoblasts, all the detectable TDP-43 was located in the nucleus in patient- and control-derived myoblasts, including in cells carrying a *TARDBP* mutation (fALS_1 patient) (Supplementary Fig. 3a), and while some FUS was found in cytoplasmic granules, these were of equal abundance in CTRL and ALS myoblasts (Supplementary Fig. 3b). Thus, the only discernible differences in TDP43 and FUS were protein abundance.

Overall, these data reflect that muscle is intrinsically affected in ALS, independently of the inputs from MNs and other environmental signals, and that downregulation of TDP-43 and FUS may be contributing to this.

### Dynamic expression of TDP-43 and FUS is required for myogenesis

To further understand the implication of TDP-43 and FUS in ALS-related myogenic alterations, we knocked these genes down in an established human myoblast cell line (8220) to further analyse changes in the myogenic capacity as well as the underlying mechanisms (Fig. 2a). Lentiviral delivered shRNAs to TDP-43 or FUS decreased expression of the target gene by 80% after 2 days (Fig. 2b). *TARDBP* silencing reduced FUS levels (40%) as well (Fig. 2b), implying an upstream regulation of TDP-43 on FUS expression. This result aligns with previous CLIP-seq studies that identified *FUS* mRNA as a target of TDP-43 [56]. Both TDP-43- and FUS-silenced myoblasts displayed ultrastructure abnormalities revealed by transmission electron microscopy (TEM) images (Supplementary Fig. 4a). Compared to scramble RNA-transduced (shRNA) myoblasts, shTDP-43 myoblasts showed a rounded and enlarged shape accompanied by mildly expanded nuclei. Organelles were arranged in perinuclear clusters, containing very short rough endoplasmic reticulum (RER) but a high number of smaller mitochondria (Supplementary Fig. 4a, b). Similarly, shFUS myoblasts were elongated with abundant mitochondria and dilated RER (Supplementary Fig. 4a, b). Despite these changes in cellular morphology and distribution of intracellular components, all silenced myoblasts were viable.

**Figure 2.**
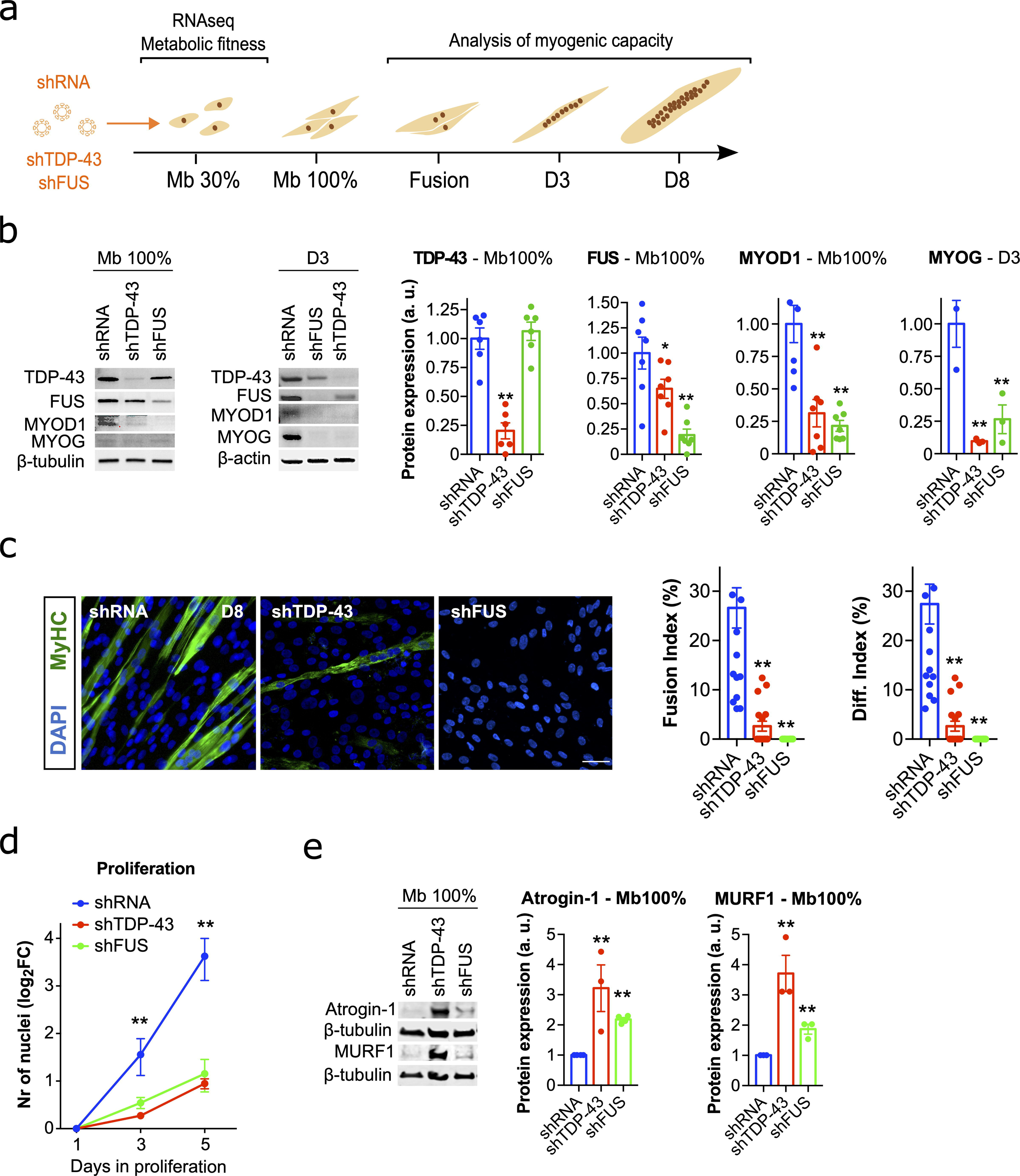
Functional consequences of TDP-43 or FUS silencing in the immortalised human control myoblasts. **a** Schematic diagram of the myogenic process showing the different phases under study, as well as the interventions and measures performed. **b** Representative Western blot images of TDP-43, FUS, MYOD1 and MYOG in shTDP-43 or shFUS-silenced myoblasts at 100% confluence (Mb 100%) and at 3 days after fusion (D3), and scatter dot-plots with bar graphs showing the quantification of protein levels. n=3-6 transductions per group. **c** Representative images of human immortalised myoblasts stained for MyHC (Myosin heavy chain, green) and DAPI (nuclei, blue), and quantitative scatter plots with bar graphs showing fusion and differentiation indices. Immunofluorescence was performed 8 days (D8) after the first fusion events of shRNA cells. Scale bar: 100 μm. n=6-11 images per group. **d** Proliferation curve measured as the fold-change in the number of DAPI-positive nuclei at days 1, 3 and 5 after cell seeding. n= 4-5 images per group. **e** Representative Western blot images of Atrogin-1 and MURF1 in TDP-43- or shFUS-silenced myoblasts at Mb 100%, and scatter plots with bar graphs showing the quantification of protein levels. n=3 infections per group. Bar graphs represent mean ± SEM. \**p*<0.05; \*\**p*<0.01 compared to the shRNA group via one-way ANOVA. a.u., arbitrary units.

We then studied the effect of TDP-43 or FUS silencing on myogenesis. First, we analysed the expression of the myogenic factors MYOD and MYOG, which in most patient-derived primary myoblasts and myotubes appeared downregulated (Supplementary Fig. 2a, b). Indeed, shTDP-43 and shFUS myogenic cells displayed a dramatic decrease in the total levels of both factors (Fig. 2b). Both silenced myoblasts were more sensitive to growth factor deprivation, so when differentiation was induced in such a way, shTDP-43 and shFUS myoblasts were not able to survive the differentiation challenge, implying an extreme susceptibility to starvation. For this reason, we switched to a more physiological myogenesis model, allowing spontaneous differentiation and fusion by cell-to-cell contact once myoblasts reach 100% confluence. In this scenario, control myoblasts differentiated and formed multinucleated myotubes (positive for myosin heavy chain, MyHC) three days after reaching 100% confluence. In contrast, TDP-43 and FUS knockdown displayed a dramatic impairment in differentiation and fusion, with barely any myotube formation (Fig. 2c). Furthermore, proliferation assays showed that the proliferative capacity of shTDP-43 and shFUS myoblasts was strongly reduced as well (Fig. 2d). Finally, as in patient-derived myoblasts (Supplementary Fig. 2e), we confirmed that the pro-atrophic proteins Atrogin-1 and MURF1 were significantly increased in silenced myoblasts, particularly in shTDP-43 myoblasts (Fig. 2e). Together, these results confirm an essential function of TDP-43 and FUS on myogenic differentiation and regeneration process.

It has been previously shown in mouse muscle progenitors that the nuclear exit of TDP-43 to form cytoplasmic granules in early myotubes is required for muscle differentiation and regeneration [81]. Thus, we performed different analyses to characterise TDP-43 and FUS localization patterns along the human myogenic process. We studied the immortalised human myoblast cell line 8220 [29] at five different stages: 30% confluence (proliferation); 100% confluence; fusion (myocytes); and early phases of differentiation until myotube formation (days D3 and D8 post-fusion) (Supplementary Fig. 5a). The analysis of the expression of the myogenic regulatory factors MYOD and MYOG showed an increase on MYOD levels prior to first fusion events followed by a decrease during fusion and myotube maturation, while MYOG appeared increased during fusion events and early phases of myotube formation (Supplementary Fig. 5b), as previously reported in rodent and human myogenesis [47, 62]. Regarding TDP-43 and FUS localization across myogenesis, TDP-43 appears mainly limited to the nucleus throughout the myogenic process, with a slighter increment in later stages, whereas nuclear FUS decreases throughout differentiation, being totally absent in early myotubes at D8 (Supplementary Fig. 5c, d). Although there was no cytoplasmic TDP-43 in myoblasts, TDP-43 appeared in cytoplasmic puncta in differentiating myotubes at D8 (Supplementary Fig. 5d), in agreement with the previous study of Vogler and collaborators showing cytoplasmic translocation of TDP-43 into RNA myogranules at this stage [81]. Together, our data confirm changes in the subcellular localization of both TDP-43 and FUS along myogenesis, suggesting their involvement in the regulation of gene expression programs along muscle differentiation.

### TDP-43 or FUS deficiency curtails anaerobic glucose metabolism and induces pro-atrophic pathways

Myogenesis poses a metabolic challenge for cells as the reliance on different metabolic pathways changes throughout the differentiation process to meet specific energy demands. Moreover, disturbances in energy metabolism have been repeatedly associated with the pathogenesis of ALS [91] and TDP-43 is a recognized regulator of skeletal muscle glucose homeostasis [9, 75]. Thus, we first investigated the extent of the metabolic reconfigurations that occur in control myoblasts during myogenesis (Supplementary Fig. 6). Metabolic flux analysis confirmed that glycolysis was the predominant metabolic pathway to produce basal ATP for myoblast proliferation, as indicated by higher extracellular acidification rates (ECAR) and lower oxygen consumption rates (OCR) (Supplementary Fig. 6a-c). However, differentiating and fusing myocytes as well as early myotubes switch their metabolism from glycolytic ATP production to oxidative phosphorylation (Supplementary Fig. 6a-c). To assess the impact of changes in metabolism, we treated proliferating myoblasts with 2-deoxyglucose (2-DG) to reduce glycolysis-dependent ATP production. 2-DG induced a myogenesis blockade (Fig. 3a, b), and decreased MYOD and MYOG levels followed by an increase in Atrogin-1 (Fig. 3c-e).

**Figure 3.**
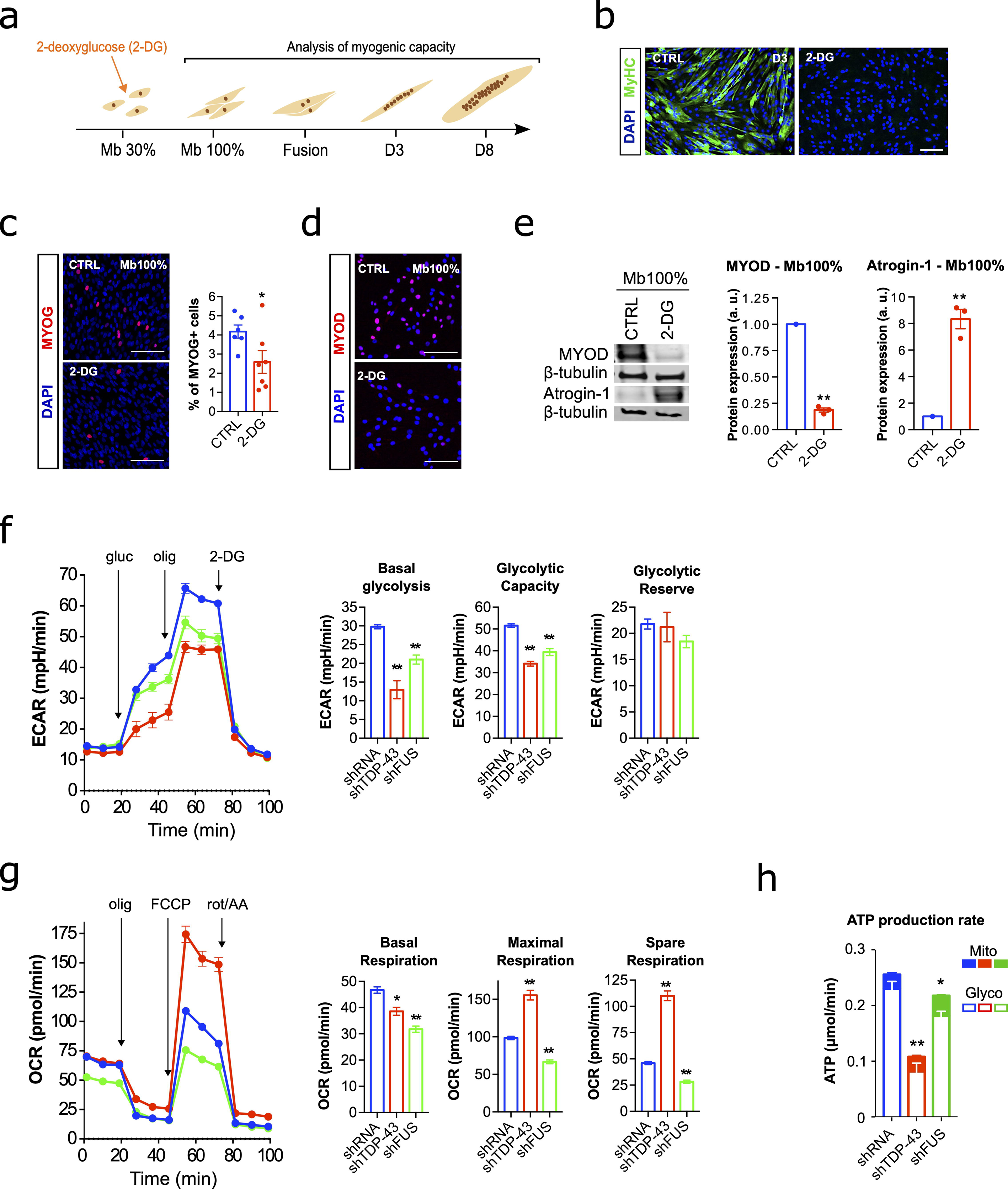
Metabolic alterations of TDP-43 or FUS silencing in the immortalised human control myoblasts. **a** Schematic diagram of the myogenic process illustrating the different phases under study, as well as the intervention point for glycolysis repression and the measures performed throughout the experiment. **b** Representative images of myogenic differentiation following 2-deoxyglucose (2-DG) treatment at day 3 after fusion (D3). Myoblasts were stained for Myosin Heavy Chain (MyHC, green) and DAPI (nuclei, blue). Scale bar: 100 μm. **c** MYOG protein expression after 2-DG treatment at 100% confluence (Mb 100%). Representative images of myoblasts stained for MYOG (red) and DAPI (blue). Scatter plots with bar graphs quantify the percentage of MYOG-positive nuclei. ImageJ software (v2.9) was used to quantify the images, with DAPI staining defining the nuclear boundaries. A nucleus was considered positive for MYOG if its staining intensity was greater than the mean +1*SD of the control (CTRL) values. n = 6-7 images per group. Scale bar: 100 μm. **d** MYOD protein expression after 2-DG treatment at Mb 100%. Representative images of myoblasts stained for MYOD (red) and DAPI (blue). n = 6-7 images per group. Scale bar: 100 μm. **e** Representative Western blot images of MYOD and Atrogin-1 protein levels, and scatter dot-plots with bar graphs showing the quantification of protein levels. Myoblasts were treated with 2-DG at Mb 100%. n = 3 treatments per group. Data are expressed as mean ± SEM. \**p*<0.05; \*\**p*<0.01 compared to the Mb 100% group via Mann-Whitney *U* test. a.u., arbitrary units; CTRL, untreated control. **f-h** Graphs for Extracellular Acidification Rate (ECAR) (**f**), Oxygen Consumption Rate OCR (**g**), and ATP production rate (**h**) of silenced myoblasts (shRNA, shTDP-43 or shFUS). The corresponding quantification of basal glycolysis, glycolytic capacity, glycolytic reserve, basal respiration, maximal respiration, and spare respiration are presented on the right side of panels (**f**) and (**g**), respectively. Analyses were performed one day after seeding myoblasts at Mb 100%. n = 4, 3, and 1 independent experiments, respectively. gluc, Glucose; olig, Oligomycin; FCCP, Carbonyl cyanide p-(trifluoromethoxy) phenylhydrazone; rot/AA, Rotenone/Antimycin A. Bar graphs represent mean ± SEM. \**p*<0.05; \*\**p*<0.01 compared to the shRNA group via one-way ANOVA.

Thus, we sought to investigate the metabolic effects of TDP-43/FUS knockdown on human myoblasts, and whether the myogenic defects induced by loss of TDP-43/FUS may be explained by glucose metabolism abnormalities. We found that basal glycolysis, maximal glycolytic capacity and glycolysis-derived ATP production, as calculated from ECAR values, were consistently repressed after silencing TDP-43 or FUS (Fig. 3f). The knockdown of FUS, and to a lesser extent of TDP-43, also affected mitochondrial respiration, as observed by a reduction in basal OCR and mitochondrial-derived ATP production (Fig. 3g, h). However, the maximal and spare respiratory capacity was found increased in TDP-43-silenced myoblasts (Fig. 3g), although this could be due to the higher number of mitochondria (Supplementary Fig. 4 a, b) that would respond under experimental conditions to the uncoupler of mitochondrial oxidative phosphorylation FCCP.

These findings suggest that both TDP-43 and FUS silencing resulted in overall abnormalities in energy metabolism during myogenic differentiation. However, the impact was particularly pronounced in the glycolytic flux, linking disturbances in glycolytic metabolism with myogenic deficiencies in silenced myoblasts. Thus, collectively, these data suggest that glucose energy homeostasis may be a key factor underlying the effects that TDP-43 or FUS silencing have on muscle formation, essentially by impairing myogenesis but also by inducing pro-atrophic programs.

### TDP-43 and FUS silencing dysregulates FOXO transcription factors and related signalling pathways

Since TDP-43 and FUS LoF are associated with extensive changes in gene expression and splicing events [26], we employed RNA sequencing to define the global gene expression profiles of silenced myoblasts in an attempt to gain insights into the pathways and processes modulated by TDP-43 and FUS that might be relevant for muscle homeostasis. Overall, 1,468 and 357 genes were upregulated (log_2_ fold change ≥1, false discovery rate [FDR]≤ 0.001), whereas 1,597 and 883 genes were downregulated (log_2_ fold change ≤ −1, FDR ≤ 0.001) (Supplementary Table 2) in shTDP-43 and shFUS cells, respectively. Almost two-thirds of the differentially expressed genes in shFUS myoblasts overlapped those in shTDP-43 myoblasts (62%; 767 out of 1,240); representation factor: 4.03; *p*<0.001) (Fig. 4a), of which 95% (730 out of 767) had the same pattern of dysregulation and similar degree of fold change (Fig. 4b). To characterise the functional effects of TDP-43 or FUS scarcity in myoblasts, we carried out GSEA using the Hallmark Pathway. Interestingly, we found consistent gene sets enrichment in myogenesis with negative gene regulation across TDP-43 and FUS-silenced myoblasts (Fig. 4c; Supplementary Table 3). Enrichment using the GO biological process, GO molecular function and KEGG pathways with GSEA confirmed the significant enrichment with negative gene regulation of sarcomere organisation, muscle organ development, striated muscle contraction and striated muscle cell differentiation among others (Supplementary Fig. 7a-c and Supplementary Table 3). Furthermore, the Hallmark GSEA also revealed significant enrichment with negative gene regulation of Myc-related targets and MTORC1 signalling gene sets (Fig. 4c), which can contribute to the dysregulation of the fine tuning between metabolism and cell differentiation during myogenesis [63]. To delineate the transcriptional regulators controlling these altered gene programs of muscle development/differentiation, we performed a motif-centric transcription factors (TF) activity analysis by ISMARA (Integrated System for Motif Activity Response Analysis [1]) to characterise the TFs whose motifs’ activities most significantly change by TDP-43 or FUS knockdown. This analysis identified a deactivation across shTDP-43 and shFUS-silenced myoblasts of transcriptional regulators related to the functional programs identified by GSEA, myogenesis and muscle contraction, including MEF2A, MEF2C, MEF2D, MYOD1 and MYOG (Fig. 4d and Supplementary Table 4); these last two are in agreement with the substantial decreases in their protein levels (Fig. 2b). Thus, our robust transcriptomic analysis reflects the molecular and functional experimental phenotypes, thus offering reliability to any mechanistic predictions obtained from this dataset. In this line, we sought candidates among the top transcriptional regulators (identified by ISMARA) with consistent activation across shTDP-43 and shFUS-silenced myoblasts for abnormal transcriptional activity related to the associated phenotypes (Fig. 4d and Supplementary Table 4). The enrichment of FOXO1/FOXO6 TF motifs was expected because myogenesis and muscle growth require the repression of muscle atrophy programs regulated by FOXO1 [22, 68], and because transcriptional activity of FOXO1 can be pharmacologically modulated [43]. Moreover, FOXO TFs are also implicated in cell metabolism, and they integrate insulin signalling with glucose and lipid metabolism to orchestrate cellular processes such as cell differentiation and autophagy [31, 85]. Thus, we next sought to explore the link between the family of FOXO TFs (focusing on FOXO1 and FOXO3 due to their involvement in the skeletal muscle) and the two main functional phenotypes observed after TDP-43 and FUS silencing: the defects in myogenic differentiation, including upregulation of pro-atrophic pathways, and metabolism.

**Figure 4.**
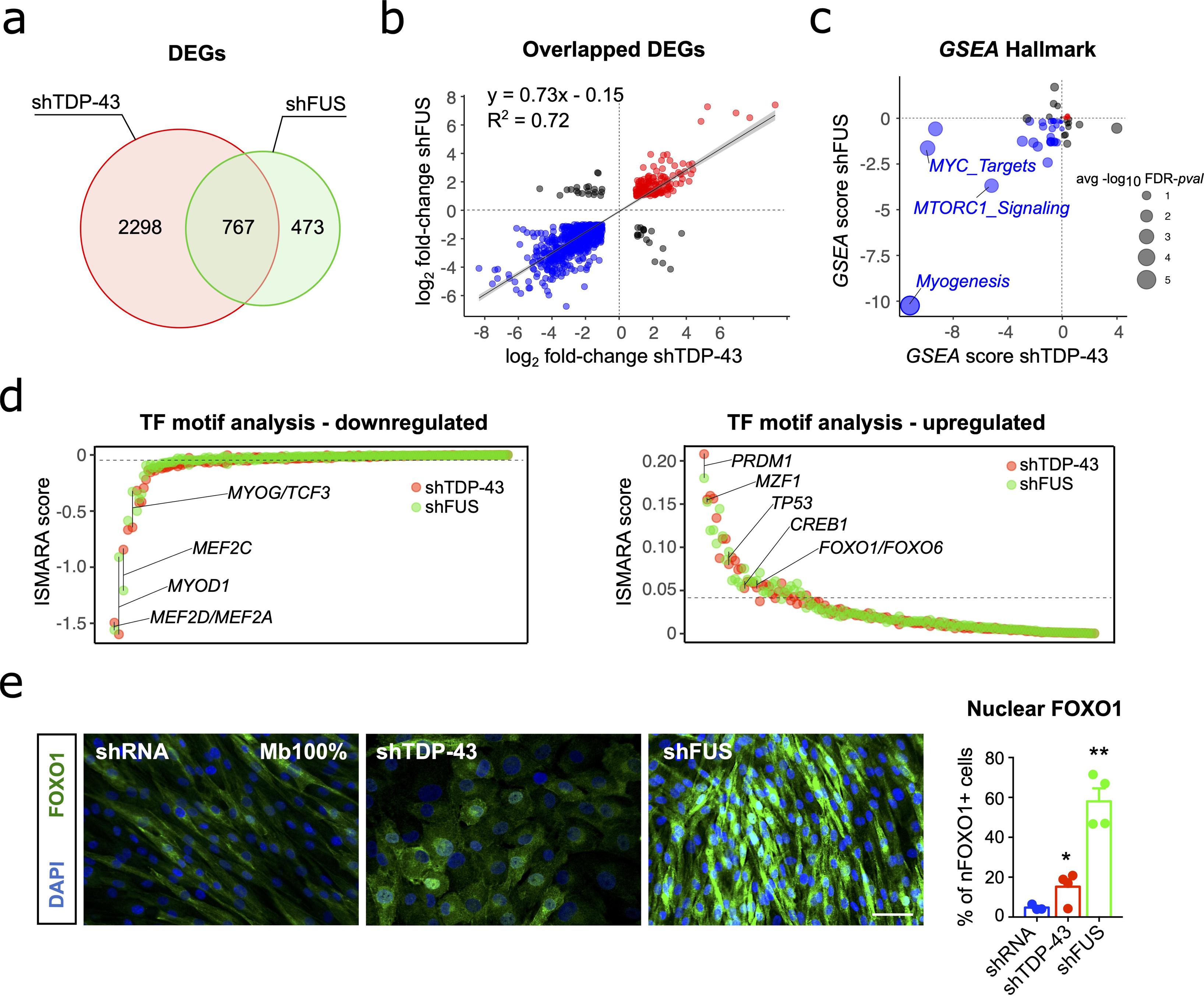
FOXO transcription factors and related signalling pathways are dysregulated upon TDP-43 or FUS silencing. **a** Venn diagram illustrates the number of overlapped differentially expressed genes (DEGs) between the shTDP-43 and the shFUS-silenced myoblasts at 100% confluence. **b** Scatterplot for log_2_-transformed fold changes of the overlapping DEGs, showing a significant correlation between shTDP-43 and shFUS-silenced myoblasts. R^2^ and linear function are indicated in the graph. Genes upregulated and downregulated in both datasets are indicated in red and blue, respectively. The grey shade area represents the 95% confidence interval. **c** Bubble chart of ranked enrichment analysis of shTDP-43 plotted against shFUS gene expression signatures using the Hallmark database by the GSEA multilevel enrichment test; GSEA score rank is based on normalised enrichment score (NES) and −log_10_FDR *p*-value for each gene set. Bubble size represents the average −log_10_FDR *p*-value. Gene sets upregulated and downregulated in both shTDP-43 and shFUS myoblasts are indicated in red and blue, respectively. Enrichment analysis of gene expression signatures using KEGG, GO Biological Process and GO Molecular Function can be viewed in *Supplementary Fig. 7*. **d** Common transcription factors (TF) across the shTDP-43 and shFUS myoblasts with predicted significant activity change by ISMARA motif analysis (based on Z score and average gene target expression change). MYOG/TCF3, MYOD1 and MEF2A/C/D are highlighted among the TF motifs with downregulated target gene expression change (left panel). FOXO1/6, CREB1, TP53, MZF1 and PRDM1 are highlighted among the TF motifs with upregulated target gene expression change (right panel). **e** Representative images of human control immortalised myoblasts at 100% confluence stained for FOXO1 (green) and DAPI (nuclei, blue), and scatter dot-plot with a bar graph showing the percentage of cells positive for nuclear FOXO1 staining. Quantification was made with ImageJ v2.9 software using DAPI staining to delimit nuclei. Nuclei with integrated density values above the cut-off point (set as the value of mean +1*SD of shRNA cells) were considered positive for FOXO1. Scale bars: 50 μm. n=6-9 images per cell line. Scale bar: 25 μm. n=3-4 images per group, 2 independent experiments. \**p*<0.05, \*\**p*<0.01 compared to the shRNA group via one-way ANOVA.

FOXO1 is particularly identified as key negative regulator of muscle growth, metabolism, and differentiation but is crucial for myoblast fusion [4, 46, 85]. Specifically, FOXO1 activation in muscle cells upregulates Atrogin-1 and MURF1 genes and downregulates MYOD expression [27, 39, 40], which mimics the effects of TDP-43 or FUS knockdown in myoblasts (Fig. 2b, e). Since transcriptional activation of FOXO requires its translocation to the nucleus, we analysed and quantified the nuclear localization of FOXO proteins across the myogenic process as well as after TDP-43 or FUS silencing in myoblasts. In control conditions, nuclear expression of FOXO1 in proliferative myoblasts was low, but it increased during the fusion events, with a concomitant decrease in cytoplasmic expression in early myotubes, which suggests translocation to the nucleus (Supplementary Fig. 8a). This result agrees with previous findings showing that FOXO1 impedes myogenic re-activation in myotubes [49]. However, nuclear FOXO1 became abnormally elevated in shTDP-43 and shFUS proliferating myoblasts, indicated by a rise in the percentage of cells with FOXO1-positive nuclei compared to shRNA cells (3.19-fold and 12.14-fold change, respectively) (Fig. 4e)., corroborating the activation of the FOXO1 TF motif inferred by ISMARA analysis. To investigate whether glycolytic repression activates FOXO1, we treated control myoblasts with 2-DG. This resulted in an increase in the number of cells with nuclear FOXO1 (8.26-fold), comparable to the effect observed upon FUS silencing (Supplementary Fig. 8b and Fig. 4e). Thus, the 2-DG-induced changes in MYOD and Atrogin-1 (Fig. 3d, e) are likely mediated by FOXO1 activation, and equally, FOXO1 activation after TDP-43/FUS silencing can be attributed to glycolytic repression.

In contrast to FOXO1, FOXO3, crucial for proliferation, exhibited a predominantly nuclear localization in control myoblasts (Supplementary Fig. 8a) as previously described in murine myoblasts [22], with decreased levels during differentiation (Supplementary Fig. 8a). However, levels of nuclear FOXO3 were also increased in shTDP-43 cells (2.86-fold) and in shFUS cells (2.32-fold) when compared to shRNA-proliferating myoblasts (Supplementary Fig. 9a).

Together, our data point to an early activation of FOXO1 activity in myoblasts with TDP-43 or FUS silencing, most likely related to defects in glucose utilization pathways with deleterious consequences for myogenesis.

### FOXO1 inhibition rescues myogenic and metabolic defects in ALS myoblasts

We next tested whether FOXO1 inhibition rescues the underlying effects of TDP-43/FUS deficiency in myoblasts. Targeting FOXO1 mRNA with shRNAs (shFOXO) reduced FOXO1 levels in TDP-43 and FUS-knockdown myoblasts (Supplementary Fig. 10), and improved their myogenic capacity as evidenced by increased differentiation and fusion indices (Fig. 5a). To elucidate further the role of FOXO1, we employed AS1842856 (iFOXO1), a quinolone-derived molecule that selectively inhibits FOXO1 transactivation. With 100-fold greater potency in targeting FOXO1 DNA interaction than other FOXO factors, iFOXO1 effectively disrupted FOXO1-mediated transactivation [43]. Strikingly, iFOXO1 treatment (33 nM) significantly improved the myogenic capacity of TDP-43-silenced myoblasts. This was evidenced by a increase in the differentiation index (4.08-fold) and in the fusion index (6.59-fold) in shTDP-43 myoblasts. Furthermore, iFOXO1 treatment activated myogenesis in shFUS-silenced myoblasts (Fig. 5b). These effects were similar to those observed in shFOXO1 knockdown (Fig. 5b). In addition, iFOXO1 treatment led to a significant increase in MYOD1 protein levels in shTDP-43 myoblasts (2.01-fold) and in shFUS myoblasts (3.02-fold) (Fig. 5c). Finally, as muscle dysfunction of silenced myoblasts was accompanied by altered energy metabolism (Fig. 3f), we also analysed the effect of FOXO1 inhibition in this context. Indeed, iFOXO1 rescued the glycolytic flux impairment caused by TDP-43 or FUS silencing, resulting in a significant increase in basal glycolysis (1.61-fold and 1.67-fold, respectively) (Fig. 5d). In addition, iFOXO1 treatment also improved the mitochondrial ATP production (Fig. 5d), thus, exerting thorough benefits in the pathways of energy production in the absence of TDP-43 or FUS.

**Figure 5.**
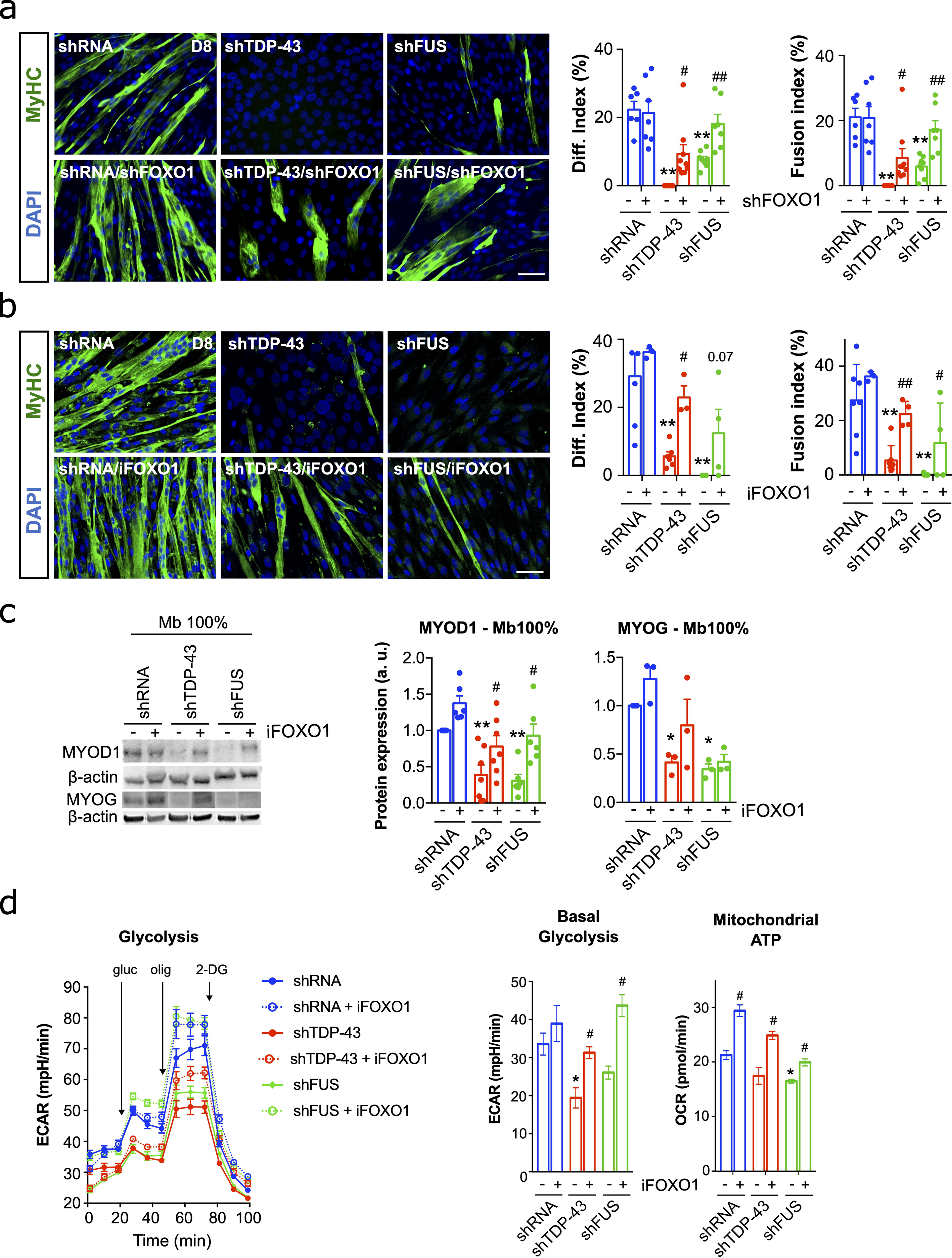
FOXO1 inhibition corrects myogenic defects and metabolic derangements induced by deficiency of TDP-43 or FUS. **a** Representative images of TDP-43 or FUS-knockdown myoblasts treated or not with shFOXO1 lentiviral particles and stained for MyHC (Myosin heavy chain, green) and DAPI (nuclei, blue). Quantitative scatter plots with bar graphs showing differentiation and fusion indices (right). Scale bar: 50 μm. n=5-6 images per group. **b** Representative images of TDP-43 or FUS-knockdown myoblasts treated or not with the selective FOXO1 inhibitor AS1842856 (iFOXO1) at 30 nM and stained for MyHC (green) and DAPI (blue). Quantitative scatter plots with bar graphs showing differentiation and fusion indices (right). Immunofluorescence was performed 8 days (D8) after the first fusion events of shRNA control cells. Scale bar: 50 μm. n=5-6 images per group. **c** Representative Western blot images of MYOD1 and MYOG protein levels in shTDP-43 or shFUS-silenced myoblasts at 100% confluence (Mb 100%) treated with iFOXO, and scatter dot-plots with bar graphs showing the quantification of protein levels. n=3-6 transductions per group. a.u., arbitrary units. **d** Representative graphs showing Extracellular Acidification Rate (ECAR) of TDP-43 or FUS-knockdown myoblasts treated or not with the selective iFOXO1 at 30 nM, and the basal glycolysis and basal respiration calculated by the data obtained in these analyses. Analyses were performed one day after seeding myoblasts at Mb 100%. n=3 independent experiments. Bar graphs represent mean ± SEM. \**p*<0.05; \*\**p*<0.01 compared to the shRNA control group via one-way ANOVA; ^#^*p*<0.05; ^##^*p*<0.01 compared to the non-treated group via one-way ANOVA. gluc, Glucose; olig, Oligomycin; 2-DG, 2-deoxyglucose; OCR, Oxygen Consumption Rate, FCCP, Carbonyl cyanide p-(trifluoromethoxy) phenylhydrazone; rot/AA, Rotenone/Antimycin A.

We next analysed FOXO expression in ALS patient-derived myoblasts by immunofluorescence. Nuclear FOXO1 levels were significantly elevated in ALS compared to CTRL myoblasts, evidenced by an increase in the percentage of cells with FOXO1-positive nuclei (2.85-fold) (Fig. 6a). We also observed a slight increase in the levels of nuclear FOXO3 (Supplementary Fig. 9b). Hence, ALS and TDP-43 or FUS silenced myoblasts share the same nuclear distribution of FOXO1/3 (Fig. 4e). Additionally, FOXO1 protein was also found to be increased in the cytoplasm of ALS patient-derived myoblasts compared to controls (3.26-fold) (Fig. 6a). iFOXO1 treatment specifically improved myogenic capacity in ALS myoblasts, as evidenced by the enhanced myogenic differentiation and fusion indices in ALS myoblasts. Compared to control myoblasts, ALS myoblasts exhibited a marked increase in the differentiation index (3.32-fold) and fusion index (3.47-fold). Notably, 7 out of 8 ALS myoblasts responded positively to the treatment (Fig. 6b). Conversely, CTRL myoblasts displayed a weaker increase in differentiation (1.61-fold) and fusion (1.59-fold) indices. The treatment also boosted glycolytic flux in the sALS myoblasts (Fig. 6c). Together these data suggest that FOXO1 inhibition improves myogenic and metabolic outcomes in ALS muscle cells.

**Figure 6.**
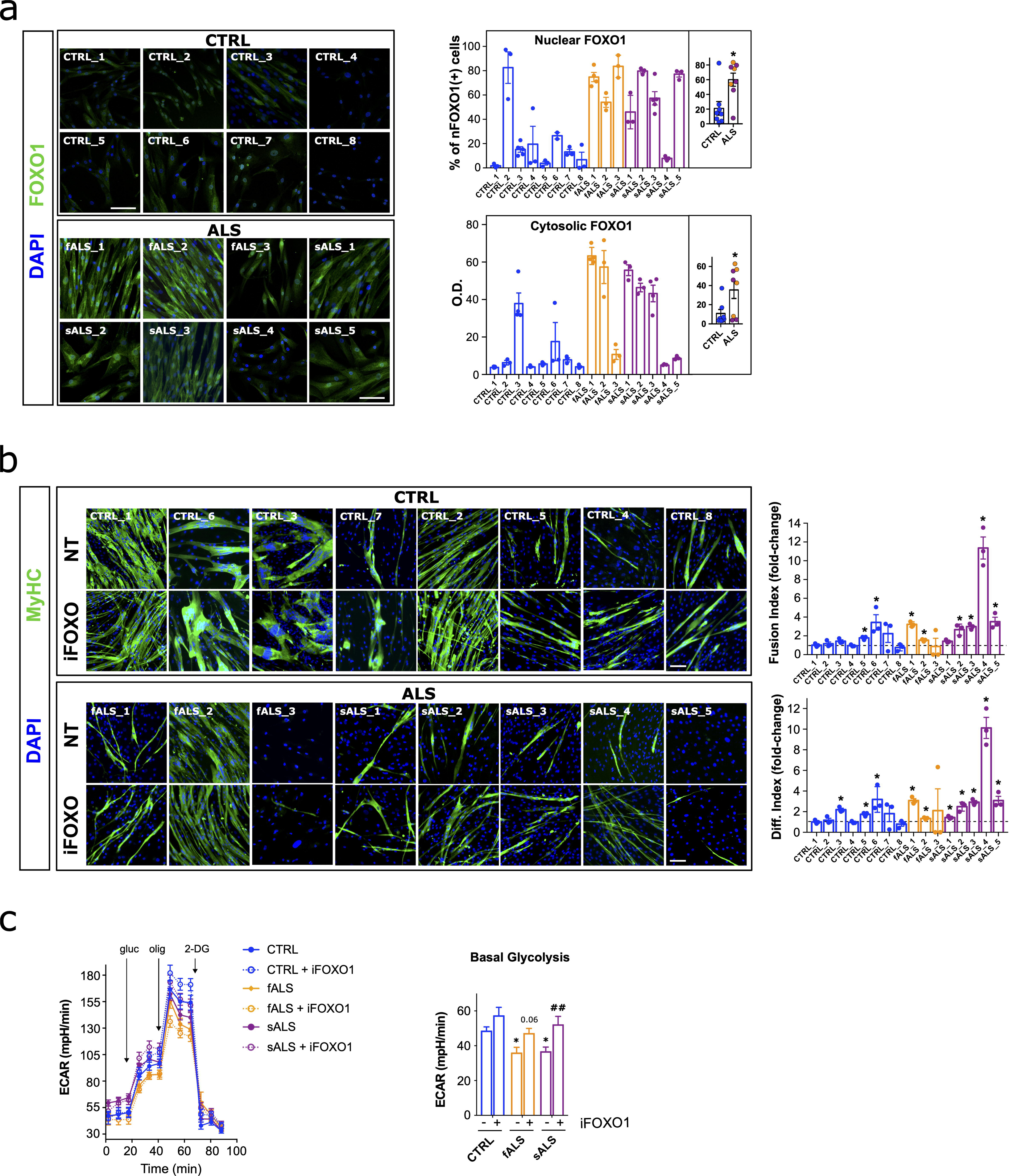
FOXO1 is dysregulated in primary myoblasts from ALS patients and its inhibition alleviates myogenic defects. **a** FOXO1 protein expression in myoblasts from ALS patients and controls. Left panel: Representative images of primary myoblasts at 100% confluence (Mb100%) from ALS patients, both familial (fALS) and sporadic ALS (sALS), and healthy controls (CTRL) stained for FOXO1 (green) and DAPI (nuclei, blue). Right panel shows a dual representation of FOXO1 staining: scatter plot showing the distribution of cells positive for nuclear FOXO1 (top), and bar graph depicting the mean optical density (OD) of cytosolic FOXO1 staining (bottom). Quantifications were done using ImageJ v2.9 software. DAPI staining was used to define nuclear boundaries. Nuclei with a staining intensity greater than the mean +1*SD of CTRL values were considered positive for FOXO1. The inner plots depict the comparison of the average values of each ALS patient group (fALS and sALS) with the CTRL group. Bar graphs represent mean ± SEM. n = 3-5 images per group. \**p*<0.05 compared to CTRL via Mann-Whitney *U* test. Scale bars: 100 μm. **b** Effect of FOXO1 inhibition on myogenic differentiation. Left panel: Representative images of primary myoblasts from ALS patients and CTRLs, treated or not (NT) with the selective FOXO1 inhibitor AS1842856 (iFOXO1) at 30 nM, stained for Myosin Heavy Chain (MyHC, green) and DAPI (nuclei, blue). Right panel: Quantitative scatter plot with bar graphs showing fusion and differentiation indices. Immunofluorescence was performed 8 days (D8) after the first fusion events observed in untreated CTRL cells. Scale bars: 100 μm. n = 5-6 images per group. Bar graphs represent mean ± SEM. \**p*<0.05 compared to the NT group via Mann-Whitney *U* test. **c** Effect of FOXO1 inhibition on glycolysis in primary myoblasts. Representative graphs show the Extracellular Acidification Rate (ECAR) of primary myoblasts from a selection of ALS patients and an age-matched healthy CTRLs, treated or not with the selective iFOXO1 at 30 nM. The basal glycolysis and glycolytic capacity were calculated based on these analyses. Analyses were performed one day after seeding myoblasts at 100% confluence. n = 3 independent experiments. Bar graphs represent mean ± SEM. \**p*<0.05, \*\**p*<0.01 compared to the NT CTRL group via one-way ANOVA; ##*p*<0.01 compared to the NT group via one-way ANOVA. gluc, Glucose; olig, Oligomycin; 2-DG, 2-deoxyglucose.

To determine whether these findings extended to other neurogenic atrophies, we studied primary myoblasts from different disorders including other MN diseases, such as SMA3 or SBMA. As an example of neuropathy, we also analysed primary myoblasts from a patient with Charcot-Marie-Tooth type 4 (CMT4; recessive and demyelinating disease) (Supplementary Fig. 11). We first examined the proliferation rates of these samples, which, collectively, were not affected in the disease group. However, at the individual level, we observed an apparent decrease in SMA3 sample (Supplementary Fig. 11a). We then compared the total protein levels of TDP-43 and FUS by immunoblotting, finding some variability, but no general pattern of change. However, SMA3 and SBMA myoblasts displayed FUS downregulation, whereas CMT4 myoblasts showed TDP-43 upregulation (Supplementary Fig. 11b). Expression of FOXO1 in these cells was similar to CTRL myoblasts, with a low nuclear percentage (<15%; Supplementary Fig. 11c), suggesting a specific pathological mechanism for ALS (65%; Fig. 6a). Finally, differentiation assays did not suggest any appreciable impairment when comparing this group of diseases with the CTRL group. However, SMA3 and, particularly, CMT4, as well as the aged CTRL (CTRL_6, 7 and 8) myoblasts, displayed a low myogenic capacity (Supplementary Fig. 11d, e). Interestingly, FOXO1 inhibition with iFOXO1 rescued the myogenic deficit only in SMA3 (Supplementary Fig. 11f).

Finally, we studied myoblasts from patients with muscular disorders characterized by protein aggregates resembling those found in ALS, such as a sporadic Inclusion Body Myositis (IBM) and a vacuolar myopathy related to Valosin-Containing Protein (VCP) mutations (devoid of ALS/FTD manifestations). While FOXO1 inhibition enhanced the myogenic program in both ALS and VCP-myopathy myoblasts (Supplementary Fig. 11f), particularly in VCP-myopathy with a 2.75-fold increase in the differentiation index, we did not observe mislocalization of FOXO1 to the nucleus (Supplementary Fig. 11c) or a negative impact on myogenesis (Supplementary Fig. 11d, e).

These results suggest that a marked increase of nuclear FOXO1 in myoblasts represents a specific ALS signature, not observed in other MN diseases such as SMA and SBMA, or neuropathies like CMT4. On the other hand, FOXO1 inhibition improved differentiation and fusion indices in diseases exhibiting FUS downregulation, including SMA type 3, SBMA, and VCP disease (Supplementary Fig. 11b). In contrast, no such rescue effect was observed in CMT4 myoblasts, which correlated with unchanged FUS expression levels (Supplementary Fig. 11b).

### Inhibition of FOXO1 improves neuromuscular function and lifespan in *Drosophila* models

Our *in vitro* findings highlight the dysregulation of FOXO1 as a shared feature of ALS patient-derived myoblasts and synthetic TDP-43 and FUS deficient myoblasts. Notably, FOXO1 inhibition improved myogenic performance and metabolic fitness in these cells. Based on these results, we aimed to assess the effects of targeting FOXO factors on muscle function *in vivo*, generating different *Drosophila melanogaster* models with conditional silencing of either Cabeza (*caz*) or *TBPH* genes (human *FUS* or *TARDBP* orthologs, respectively) by dsRNA specifically in muscle tissue (Fig. 7a and Supplementary Fig. 12a). *caz* and *TBPH* knockdown were conditioned to the expression of the Myocyte enhancer factor 2 (*DmeI/Mef2*) gene, a TF whose expression is mostly restricted to both muscle differentiated and muscle precursor cells and which is essential for myogenesis [5, 56]. With this approach, we silenced *caz* or *TBPH* specifically in the skeletal muscle of *Drosophila* flies using different long-hairpin iRNA libraries (Supplementary Fig. 12a). Each generated fly model silenced a single and different locus of *caz* or *TBPH* genes. Respectively, *caz* or *TBPH* gene expression is significantly reduced in iCaz or iTBPH models (Supplementary Fig. 12b). Notably, fly models from KK libraries (iCaz and iTBPH^pkk(108354)^) silenced more efficiently *caz* or *TBPH* (by ca 50%) (Supplementary Fig. 12b). Moreover, the iTBPH^pkk(108354)^ fly model showed not only a prominent *TBPH* expression decrease but also a subsequent and significant reduction of *caz* gene expression (Supplementary Fig. 12b), as we showed for FUS in TDP-43-silenced myoblasts (Fig. 2b). This result confirms the functional co-regulation between *TBPH* and *caz* gene expression [82], as it also occurs in human orthologs TDP-43 and FUS [56].

**Figure 7.**
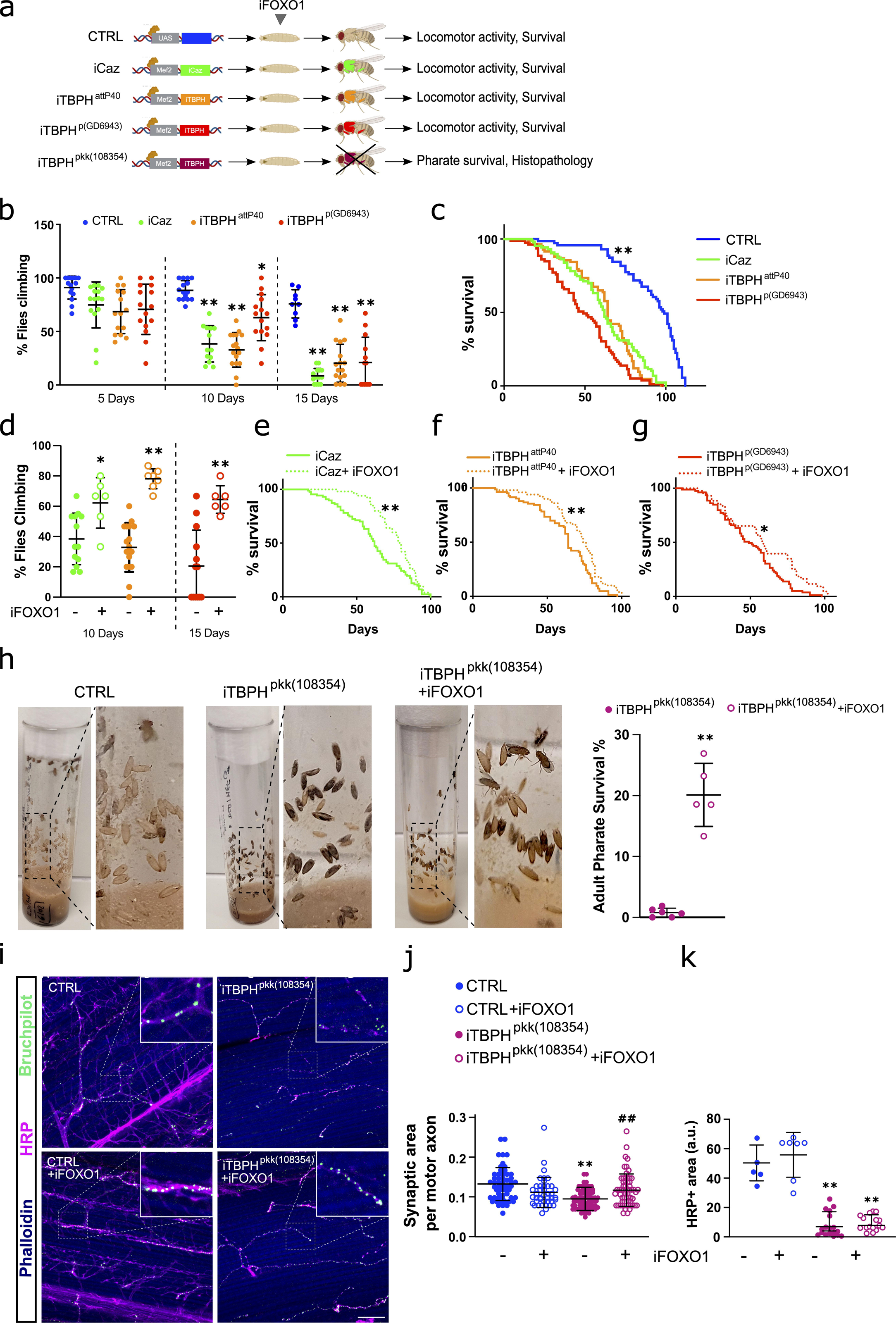
FOXO1 inhibitor AS1842856 treatment reverses phenotypical-functional parameters in *Drosophila melanogaster* models with muscle-conditioned TDP-43/FUS downregulation. **a** Schematic illustration of the different fly models used in this study and the analyses performed. Flies were orally treated (feeding) with the FOXO1 inhibitor AS1842856 (iFOXO1) from the larval stage onwards. b Scatter dot-plots with median and interquartile range showing climbing activity of the different fly models on adult days 5, 10 and 15. n= 5 flies per tube, 10 tubes per group. Each value is calculated as the average of three trials. \**p*<0.05; \*\**p*<0.01 compared to control (CTRL) flies via one-way ANOVA. c Kaplan-Meier curves of *caz-* or *TBPH*-silenced flies. n=100 flies per group; \*\**p*<0.01 compared to any of the knockdown flies via log-rank test. d Scatter dot-plots with median and interquartile range showing climbing activity of the different fly models treated with iFOXO1 at 30 µM. n= 5 flies per tube, 9 tubes per group. Each value is calculated as the average of three trials. \**p*<0.05; \*\**p*<0.01 compared to non-treated flies via two-way ANOVA. e-g Kaplan-Meier curves of *caz-* (e) or *TBPH*-silenced flies (f-g) treated (feeding) with iFOXO1. n=90 flies per group. \**p*<0.05; \*\**p*<0.01 compared to non-treated flies via log-rank test. h Representative images of pharate lethality in the fly model *iTBPH^pkk(108354)^* with or without treatment (feeding) with iFOXO1. Note that the pupae of CTRL flies are empty and adult flies are at the top of the tube. However, adult pharates of the *iTBPH^pkk(108354)^* model remain trapped inside the pupa. The scatter dot-plots with median and interquartile range in the right shows the effect of iFOXO1 on adult pharate survival. n= 75 pupae per tube, 6 tubes per group. \*\**p*<0.01 compared to non-treated flies via Student’s t test. i Representative images of *Drosophila* indirect flight muscles stained with Phalloidin (blue), neuronal marker HRP (pink) and Bruchpilot (green). Scale bar: 50 µm. Digitally zoomed images show Bruchpilot+ synaptic boutons along motoneuron terminals. j, k Scatter dot-plots with median and interquartile range shows the quantification of the synaptic area (j) and neuron (HRP+) density (k). n=12 flies per group. \*\**p*<0.01 compared to CTRL flies; ^##^*p*<0.01 compared to *iTBPH^pkk(108354)^*flies via two-way ANOVA.

While *caz*-silenced (*icaz*) flies and two different lines of *TBPH*-silencing (iTBPH^attp40^ and iTBPH^p(GD6943)^) were viable, the third TBPH-silenced line (iTBPH^pkk(108354)^) exhibited lethality in newly metamorphosed adults (pharate), not even being able to hatch out of its pupa (Fig. 7a). Adult iTBPH^attp40^ and iCaz flies exhibited the most pronounced motor phenotypes, detectable as early as 10 days old. This was evidenced by a climbing ability of only 30-40% of flies in the locomotor activity test. (Fig. 7b). However, adult iTBPH^p(GD6943)^ flies displayed such phenotype at 15-day old (Fig. 7b). Finally, to complete the phenotypical-functional characterization of the models, we performed longevity assays, demonstrating a significantly decreased lifespan across iCaz and iTBPH fly models (Fig. 7c).

*Drosophila* has only one ortholog for mammalian FOXO transcription factors, named *foxo*. Although the affinity of the AS1842856 compound (iFOXO1) for *Drosophila*’s *foxo* has never been tested, as FOXO binding sites for this compound are well conserved (Supplementary Fig. 12c), we first tested its efficacy in our models. After verifying that iFOXO1 (30 µM) showed no toxicity on motor function and lifespan in control flies (Supplementary Fig. 12d, e), we applied iFOXO1 treatment in *icaz* and *iTBPH* flies. Strikingly, *foxo* inhibition resulted notably improved muscle function, as evidenced by a significant increase in climbing activity. This effect was more pronounced in iTBPH flies (approximately 80% climbed) compared to iCaz flies (approximately 60% climbed) (Fig. 7d and Supplementary Videos 1-7). Furthermore, treated flies significantly increased their survival, particularly for the iCaz and iTBPH^attp40^ fly lines (Fig. 7e-g).

As mentioned above, iTBPH^pkk(108354)^ flies displayed adult pharate lethality, presumably due to insufficient muscle strength to exit the pupa once metamorphosis is complete. Noteworthily, iFOXO1 treatment significantly reduced pharate lethality in iTBPH^pkk(108354)^ flies. This was evidenced by a substantial increase in survival rate, from 2% to 20%, at the pharate pupal stage (Fig. 7h). Indeed, histopathological analysis revealed a dramatic denervation of iTBPH^pkk(108254)^ thoracic muscles, as observed by both a decrease in the number of HRP-positive neuronal branches (−7.14-fold) and in the size of neuromuscular synapses (−1.4-fold) (*bruchpilot*+, a marker of mature and functional synapses in *Drosophila*) (Fig. 7i-k), confirming a denervation phenotype in these *TBPH* muscle-silenced flies. Surprisingly, the treatment with iFOXO1 induced a significant increase in the size of the neuromuscular synapses of the remaining MNs (1.25-fold) (Fig. 7i, j), correlating with the survival increase. However, the treatment was unable to show signs of improvement in the number of terminal MN innervating the muscle (Fig. 7k).

Together, the combined data obtained from the *in vivo* models and *in vitro* experiments suggest a hopeful prospect for using FOXO1 inhibition to restore muscle homeostasis challenged by TDP-43 or FUS LoF, suggesting a potential benefit to reduce muscle wasting in ALS patients. Providing this initial evidence, these data also promote that therapies aimed at preserving muscle function could potentially offer protection to MNs.

## DISCUSSION

ALS is a complex and devastating disease that affects MNs and involves a multitude of altered processes and pathways, making the development of effective therapeutic strategies challenging [91]. Recent research has highlighted the implication of skeletal muscle in the disease, not only as a consequence but also as a potential contributor to the origin and spread of the pathology [17, 55, 70]. In this work, we have sought to provide further evidence of the molecular bases underlying the intrinsic muscle abnormalities of ALS to pinpoint mechanisms that can be targeted to improve muscle health and, ultimately, slow down the disease pathology. In this sense, we demonstrate that myogenesis, glucose metabolism and pro-atrophic pathways are affected in ALS muscle progenitor cells independently of the MN input, being the role of TDP-43 and FUS essential in these processes. Aberrant hyperactivation of FOXO transcription factors was identified as an underlying mechanism, evidenced by FOXO1 nuclear translocation and FOXO1 motif activity upregulation. We evidence that FOXO1 activity can be pharmacologically modulated to recover muscle homeostasis in ALS cells and to improve the neuromuscular function in ALS *Drosophila* models with muscle-specific TDP-43 and FUS silencing.

To decipher the molecular basis of muscle contribution to ALS pathology, we have focused our study on the capacity of muscle resident precursor cells (the myoblasts) to fulfil their compromise for muscle regeneration. The skeletal muscle has an extraordinary regenerative capacity to recover from severe damage and even atrophy under physiological conditions [59], by activating the myogenic program in muscle progenitor cells. However, ALS muscle atrophy is irreversible and follows a progressive pattern that determines the disease severity, being disabling and fatal for the patients [38]. A plausible hypothesis is that the regenerative capacity of ALS muscle is impaired, to the point of not compensating for denervation-induced atrophy. According to this hypothesis, we found defects in the process of muscle regeneration in almost all ALS samples, in agreement with previous studies [57, 69], which were further accompanied by changes in the levels of the myogenic regulatory factors (MRF) MYOD and MYOG. Initially, we elucidated whether two of the most implicated factors in ALS pathology, TDP-43 and FUS, might be involved in such myogenic defects. Indeed, we identified a faint presence of cytoplasmic TDP-43 granules as early as day 3 after human myocyte fusion in human control myotubes. Accordingly, a previous study reported that TDP-43 forms physiological cytoplasm granules in murine C2C12 myotubes at late stages of differentiation, which are critical for regulating sarcomeric protein translation and myotube maturation in mice [81] cytoplasmic TDP-43 protein granules in myoblasts from either ALS patients or control subjects. This suggests that during the early stages of myogenesis, cytoplasmic TDP-43 may play a minimal role. However, TDP-43 total levels appeared decreased in ALS myoblasts, suggesting that nuclear TDP-43 may also play a significant role in the early stages of differentiation by regulating myogenic gene programs. This hypothesis can also be extended to FUS, as ALS myoblasts demonstrate decreased FUS expression, which is normally localized primarily to the nuclei of proliferating myoblasts but is absent from differentiated myotubes. Nonetheless, we must consider some variability in myogenesis among ALS patients. For instance, in fALS2, the only patient without differentiation problems presents a familial form with slower disease progression. There is also more variability in healthy individuals, where we observed a clear correlation with aging. As this correlation is lost in ALS, we anticipate that myogenic impairments are genuinely associated with ALS pathology, thus, suggesting premature aging [52]. Since we reported general decreased TDP-43 and FUS expression in the ALS samples, we investigated the mechanisms and processes related to myogenesis impacted by TDP-43 or FUS knockdown in proliferating myoblasts.

TDP-43 or FUS knockdown was sufficient to replicate the myogenic defects observed in patientś cells. These results are in agreement with those achieved by the silencing of other ALS-causative genes. For instance, OPTN silencing delayed muscle regeneration in mice and impaired myogenic differentiation in C2C12 mouse myoblasts [72], and RNA-binding proteins whose mutations cause ALS, such as hnRNPA1, hnRNPA2B1 or MATR3, control myogenic cell fate transitions during differentiation by regulating the expression and alternative splicing of muscle-related genes [2, 32, 50, 83]. Hence, the diversity of ALS-associated genes influencing myogenic cell fate transitions highlights the potential for various triggers to cause myogenic alterations in ALS. While our knockdown model represents a simplified approach to the complex TDP-43 proteinopathy, it is important to note that the loss of TDP-43 function, particularly its role in cryptic splicing repression, is thought to precede the formation of TDP-43-positive inclusions [8]. Therefore, our silencing approach mimics an early stage of the disease, making it valuable for studying the underlying pathogenic mechanisms.

The myogenic process requires a series of consecutive steps that demand a fine-tuned regulation of metabolism and a flexible use of energy resources [15, 50], as we confirmed in the human immortalised myoblast cell line used in this work. In the earliest stages of myoblast differentiation, the reliance on glycolysis is critical, as we demonstrated after inhibiting myoblasts with 2-DG resulting in myogenic repression, whereas in later stages of myotube maturation, energy is mostly supplied by mitochondrial oxidative phosphorylation [50, 79]. Indeed, glycolysis not only provides a rapid source of energy and carbon sources to underpin myoblast growth but also acts as an epigenetic regulator of myoblast proliferation through modulation of histone acetylation [87]. As alterations in glucose metabolism are major within the overall metabolic disturbances of ALS [3, 91], it is plausible to anticipate that these may impinge on the myogenic process and, in turn, affect muscle function. Interestingly, the knockdown of TDP-43 or FUS in myoblasts dramatically reduced the ability to use glucose, and pro-atrophic pathways became consequently activated. These results fit previous studies describing the role of TDP-43 in the regulation of skeletal muscle glucose homeostasis, which involves glucose uptake facilitation mediated by the interaction between TBC1D1 and the glucose transporter GLUT4 [9, 75]. This highlights the important role that TDP-43 and FUS play in glucose oxidation pathways to regulate muscle function, avoiding the induction of pro-atrophic programs.

Through the analysis for enriched TF motifs from RNA-seq data, specific genes related to muscle differentiation and growth appeared to be deactivated, confirming the intrinsic muscle dysfunction. Among these factors, we found *MYOD* and *MYOG* (essential for myogenesis), which we confirmed to be downregulated in ALS samples, and *MEF2* TF, whose inactivation induces skeletal muscle atrophy [45, 71]. Regarding the activated genes, besides *FOXO1*, *TP53* appeared as another plausible candidate to be further studied, as its upregulation also suppresses cell growth, specifically by repressing *MYOG* in the skeletal muscle, leading to the inhibition of late differentiation markers [86]. As such, further analysis of these factors should be done to decipher their impact in ALS pathophysiology. However, based on the TF and our expertise in the molecular regulation of muscle stem cells [19], we uncovered dysregulation of FOXO transcription factors as a converging molecular mechanism linking the impaired myogenesis and the altered glycolysis in the absence of TDP-43 and FUS. Although FOXO1 must be excluded from the nucleus becoming inactive in myoblasts during myogenesis [22], we found FOXO1 expressed in the nuclei of all ALS patient myoblasts as well as in TDP-43- or FUS-silenced myoblasts, suggesting this change as a consistent pathogenic feature in ALS muscle. Indeed, such activation of nuclear FOXO1 did not appear in the other diseases, such as SMA3, SBMA, CMT4, IBM, or VCP. FOXO1 induction may occur as a cellular response to the inability to obtain energy from glucose oxidation. Although FOXO1 inhibits glycolysis *per se* by regulating PDK4 expression [16, 20, 58], and thus controlling the activity of the main glycolysis-limiting enzyme, pyruvate dehydrogenase (PDH), the fact that FOXO1 was activated after repressing glycolysis with 2-DG or after TDP-43 or FUS silencing could be explained by the role of FOXO1 in nutrient switching during glucose shortage [66]. Indeed, our results indicate that FOXO1 inhibition facilitates glucose metabolism, shown as an increased lactate production and basal mitochondrial respiration. On the other hand, TDP-43 directly represses nuclear FOXO1 transactivation [88]; therefore, a dysfunction of TDP-43 would release such repression leading to an increase in FOXO1 activity and, consequently, to the inhibition of glycolysis. Together, our data point to abnormal FOXO1 activation in ALS muscle due to TDP-43 or FUS knockdown, which ultimately impairs the muscle regenerative capacity. This FOXO1 overactivation may occur due to the release of FOXO1 repression in the absence of TDP-43, secondary to alterations in glucose metabolism, or both. Actually, the overactivation of FOXO1 in ALS muscle could explain the switch from glucose to muscle lipid utilisation described at early stages in the SOD1-G93A mouse, prior to symptoms onset and MN loss [12, 51].

Importantly, there are several studies that have underscored the critical role of FOXO TF in muscle atrophy [68]. Conversely, myostatin treatment, known for its negative regulation of muscle growth, reverses the IGF-1/PI3K/AKT hypertrophy pathway by inhibiting AKT phosphorylation, thereby elevating active FOXO1 levels and promoting the expression of atrophy-related genes [39]. Furthermore, FOXO1’s negative regulatory influence on skeletal muscle mass and type I fibre gene expression has been proposed, potentially contributing to sarcopenia, obesity, and diabetes in humans [25]. Additionally, FOXO3 activation, which leads to significant muscle atrophy via the transcription of atrophy-related genes, including crucial ubiquitin ligases [89], emerges as a potential therapeutic target for muscle wasting disorders and degenerative diseases involving autophagy [35], further supported by research indicating enhanced FOXO-dependent muscle mass loss associated with a decline in PGC1-alpha during atrophy [67]. Finally, Nakashima and Yakabe stated that when AMPK signalling pathway is enhanced by AICAR in C2C12 myotubes, it promotes myofibrillar protein degradation through increased expression of atrogin-1 and MuRF1, facilitated by elevated levels of FOXO TF in skeletal muscles [44]. Together, this data moved us to consider FOXO1 as a plausible target in which to intervene.

Considering our results, FOXO1 inhibitors may have the potential to treat TDP-43 or FUS LoF-induced muscle alterations, such as those in ALS. Indeed, FOXO inhibition induces overall locomotor and survival benefits in fruit flies with specific muscle knockdown of *TBPH* and *caz*. Actually, the four different *Drosophila* ALS models used in this study reproduce some ALS outcomes, such as shortened lifespan and impaired neuromuscular function, as described for TDP-43 knockdown in *Drosophila* [76]. One of the *TBPH* knockdown models exhibited a pharate lethality phenotype, likely resulting from the extensive denervation of thoracic muscles caused by *TBPH* knockdown, which in turn led to inadequate tractor capacity for exiting the pupa. Interestingly, this model displayed greater phenotypic severity compared to the others, potentially due to cumulative effects resulting from secondary *caz* dysregulation. The pharate lethality in this model was rescued by FOXO1 inhibition, which likely improved the neuromuscular synaptic contacts, favouring the subsequent pupal period eclosion. These findings support the recent theory of non-cell autonomous mechanisms of neurodegeneration in ALS [21] that implicate distal axonopathy as the initiating event in the disease [41], and provide compelling evidence that treatments aimed at restoring muscle function can alleviate MN neurodegeneration. However, it should be noted that *Drosophila* has only one *foxo* gene that performs the functions of all members of the human FOXO family, and the beneficial effects of the FOXO inhibitor treatment may be partially attributed to FOXO3 inhibition, which may also contribute to myoblast differentiation [74].

Overall, the restoration of the myogenic process being concomitant to glycolytic normalisation supports that metabolic defects are linked to the myogenic phenotypes of *TARDBP* and *FUS* knockdown cells. FOXO1 plays a crucial role in the interplay between metabolism and disease, as it controls many of the physiological responses to metabolic stimuli, including those mediated by insulin signalling [31]. Therefore, inhibiting FOXO1 could potentially counteract the key functional consequences of TDP-43 or FUS loss-of-function, specifically in ALS muscle cells. Additionally, recent studies have proposed FOXO transcription factors as promising therapeutic targets for neurodegenerative diseases [6, 48]. Indeed, we show the recovery of muscle differentiation through FOXO1 inhibition in other MN diseases such as SMA3 and SBMA. Thus, highlighting this mechanism and downstream targets could encourage the development of new therapeutic approaches for ALS that target skeletal muscle.

## Data Availability

All data produced in the present study are available upon reasonable request to the authors.

https://www.ncbi.nlm.nih.gov/geo/query/acc.cgi?acc=GSE227573

## ACKNOWLEDGMENTS

The authors thank Dr. Vincent Mouly and the Platform for Immortalization of Human Cells (Myology Institute) for the human immortalised 8220 cell line, and Stéphane Vasseur from the MYOBANK-AFM (Myology Institute) for the human CMT4 myoblasts. We also thank Dr. Francesco P. Marchese from the Genomics Unit Platform at University of Navarra & Centre for Applied Medical Research (CIMA) for expertise and assistance in NGS. We want to dedicate this work specially to all ALS patients and their families.

## Compliance with ethical standards

### Ethics approval and consent to participate

This study was approved by the Clinical Research Ethics Committees of the Basque Country and Donostia University Hospital (codes: ALM-EMP-2015-01, PI2016075, PI2019198) (Spain). Written informed consent was obtained from all the subjects included in the study.

### Consent for publication

All authors consented to publication of this paper.

## Author Contributions

ALM, FG-B and SA-M designed the study in collaboration with MZuf (*in vitro* and metabolic experiments) and GG (*Drosophila* experiments). MZuf, OP-M, MG-A, AE, ML, MZul, AS, GG and SA-M performed experiments with ALS patient-derived primary cultures and MZuf, OP-M, MG-A, XB, PI, JO, UF-P, MM-O, GF-E, AV-I, IJH and FG-B with 8220 immortalised human control myoblasts. MZuf, MG-A, GF-E and AA performed SeaHorse experiments. JMG-V and VH-P designed and performed *TEM* experiments. OA, LB and FG-B analysed NGS data. PI, JJP, RR-O, RF-T, JBE, MB, AL, GF-E, JR, EM, SD and ALM clinically identified and characterised patients and collected muscle samples. AJ and GG performed and analysed *Drosophila* experiments. ALM, FG-B and SA-M supervised and coordinated all work. ALM, FG-B and SA-M contributed to the preparation of the manuscript.

## SUPPLEMENTARY INFO

### Supplementary Tables

Supplementary Table 1: Primary antibodies.

Supplementary Table 2: DEGs of shTDP-43 and shFUS myoblasts from RNA-seq experiment.

Supplementary Table 3: GO and KEGG enriched pathways of shTDP-43 and shFUS myoblasts from RNA-seq experiment.

Supplementary Table 4: ISMARA TF motifs analysis.

### Supplementary Figures

Supplementary Figure 1. Haematoxylin and eosin staining for ALS and control muscle sections.

Supplementary Figure 2. Further characterization of primary myoblasts from ALS patients.

Supplementary Figure 3. Examination of cytoplasmic granules of TDP-43 and FUS in ALS primary human myoblasts.

Supplementary Figure 4. Ultrastructural characteristics of human myoblasts with TDP-43 or FUS silencing.

Supplementary Figure 5. Dynamics of TDP-43 and FUS along the myogenic process in the immortalised human myoblasts.

Supplementary Figure 6. Metabolic characterization of the immortalised human myoblasts.

Supplementary Figure 7. Extended Figure 4c. a-c

Supplementary Figure 8. Dynamics of FOXO transcription factors throughout myogenesis and metabolic challenges in human myoblasts.

Supplementary Figure 9. Levels of nuclear FOXO3 in myoblasts models of ALS.

Supplementary Figure 10. Effect of FOXO1 silencing on the levels of nuclear FOXO1 in human myoblasts silenced for TDP-43 or FUS.

Supplementary Figure 11. FOXO1 is not dysregulated in primary myoblasts from non-ALS neurogenic atrophies and myopathies.

Supplementary Figure 12. Extended Fig. 7.

### Supplementary Videos

Supplementary Video 1. Locomotor activity test in non-treated (NT) CONTROL flies

Supplementary Video 2. Locomotor activity test in non-treated (NT) iCaz flies

Supplementary Video 3. Locomotor activity test in iFOXO1-treated iCaz flies

Supplementary Video 4. Locomotor activity test in non-treated (NT) iTBPH^attp40^ flies

Supplementary Video 5. Locomotor activity test in iFOXO1-treated iTBPH^attp40^ flies

Supplementary Video 6. Locomotor activity test in non-treated (NT) iTBPH^p(GD6943)^ flies

Supplementary Video 7. Locomotor activity test in iFOXO1-treated iTBPH^p(GD6943)^ flies

