## Supplementary figures 1-12 for "Dysregulated FOXO1 activity drives skeletal muscle intrinsic dysfunction in amyotrophic lateral sclerosis"

**ACTA NEUROPATHOLOGICA**

**SUPPLEMENTARY FIGURES**

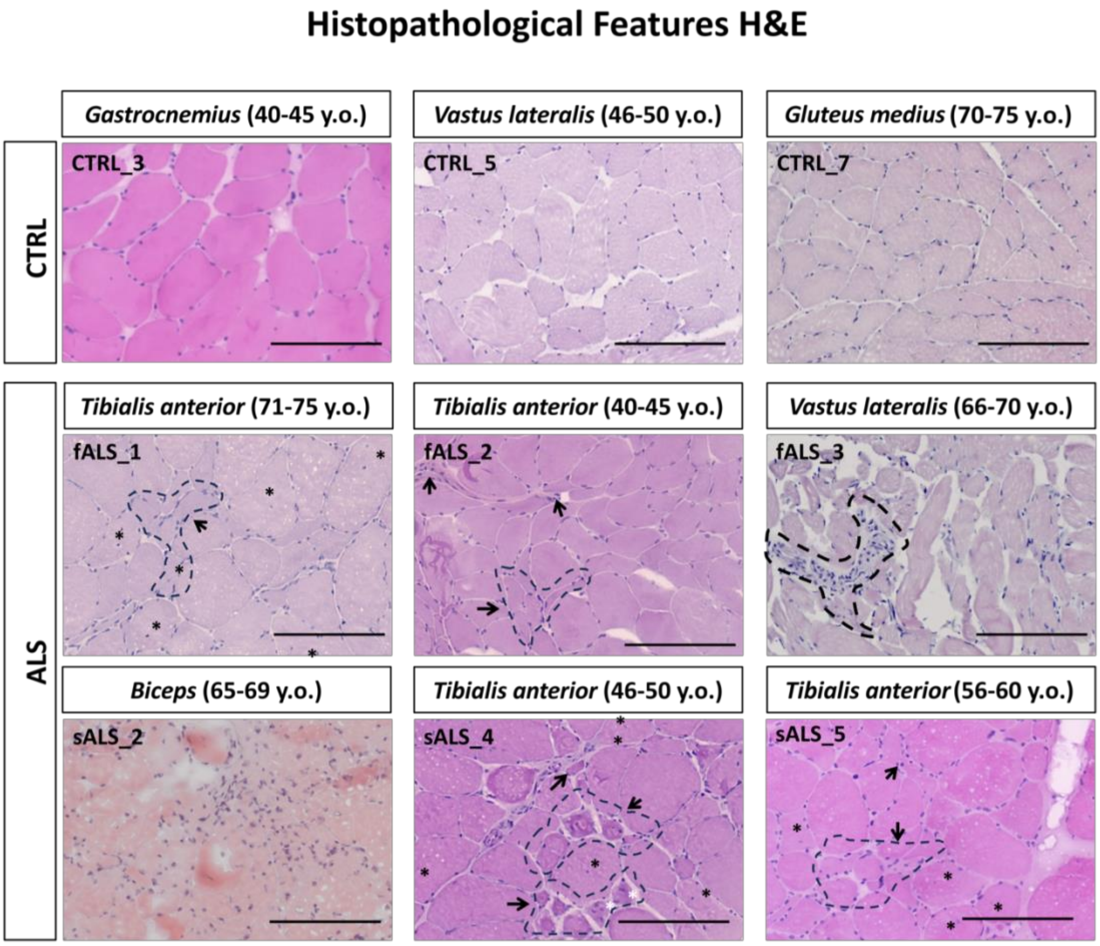


**Supplementary Figure 1. Haematoxylin and eosin staining for ALS and control muscle**. Representative images of 8 μm muscle cross-sections stained for haematoxylin and eosin from age-matched control (CTRL) and ALS patients, both familial (fALS) and sporadic ALS (sALS), as indicated. Images were scanned at 10X magnification. y.o., years old; black arrows, atrophic fibres; black asterisk, central nuclei; white asterisk, pyknotic nuclei; dotty lines; cell infiltration. Scale bar, 100 μm.

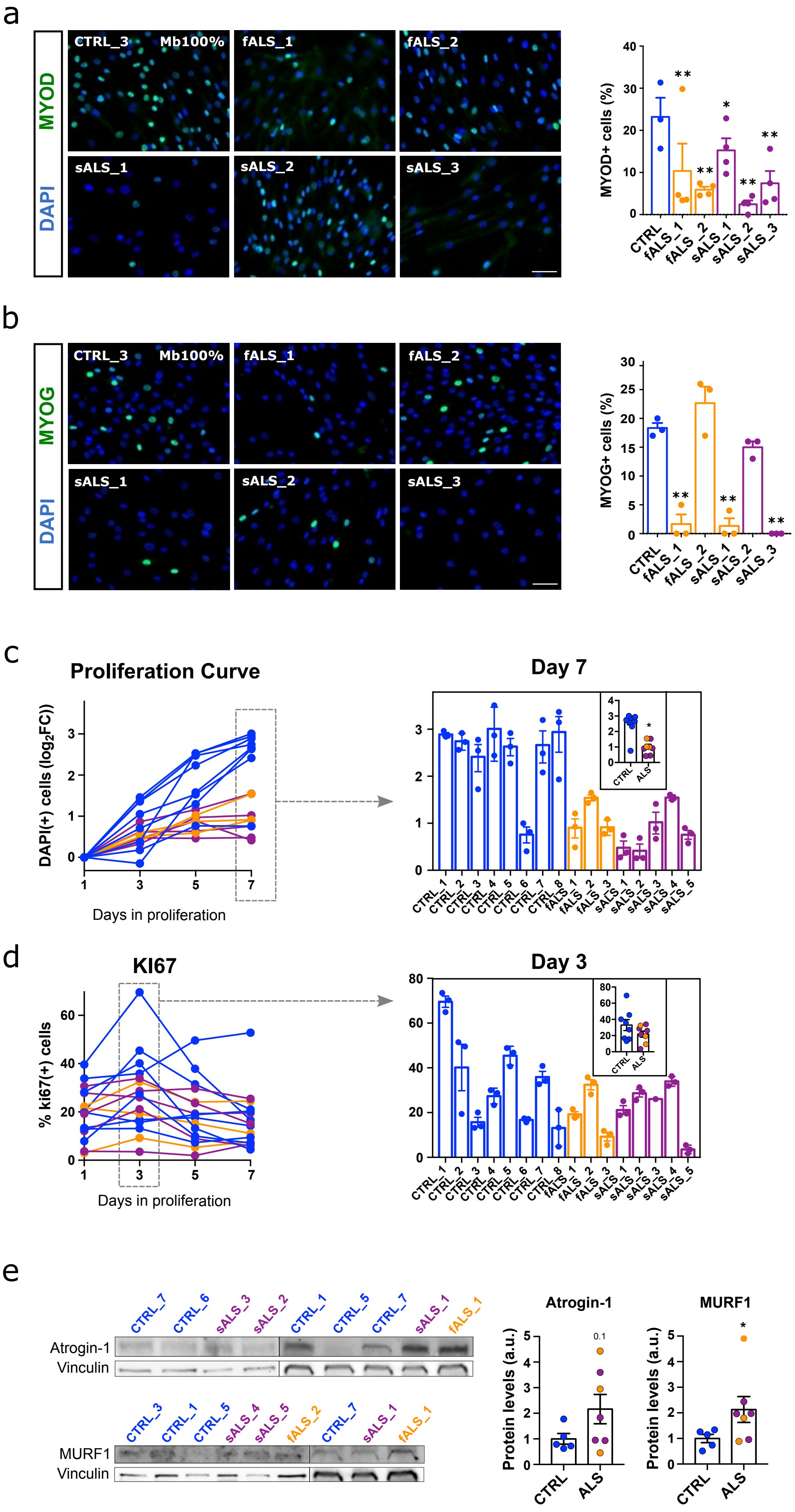

a
CTRL_3 Mb100%
fALS_1
fALS_2
sALS_1
sALS_2
sALS_3
MYOD
DAPI
MYOD+ cells (%)
CTRL fALS_1 fALS_2 sALS_1 sALS_2 sALS_3
b
CTRL_3 Mb100%
fALS_1
fALS_2
sALS_1
sALS_2
sALS_3
MYOG
DAPI
MYOG+ cells (%)
c
Proliferation Curve
DAPI(+) cells (log2FC))
Days in proliferation
Day 7
CTRL ALS
CTRL_1 CTRL_2 CTRL_3 CTRL_4 CTRL_5 CTRL_6 CTRL_7 CTRL_8 fALS_1 fALS_2 fALS_3 sALS_1 sALS_2 sALS_3 sALS_4 sALS_5
d
KI67
% ki67(+) cells
Days in proliferation
Day 3
e
CTRL_7 CTRL_6 sALS_3 sALS_2 CTRL_1 CTRL_5 CTRL_7 sALS_1 fALS_1
Atrogin-1
Vinculin
CTRL_3 CTRL_1 CTRL_5 sALS_4 sALS_5 fALS_2 CTRL_7 sALS_1 fALS_1
MURF1
Vinculin
Atrogin-1
MURF1
Protein levels (a.u.)
0.1
CTRL ALS

**Supplementary Figure 2. Further characterization of primary myoblasts from ALS patients. a-b** Left panels: Representative images of primary myoblasts from a selection of ALS patients, both familial (fALS) and sporadic ALS (sALS) and an age-matched healthy control (CTRL_3) at 100% confluence stained for MYOD (**a**) and myogenin (MYOG) (**b**). Right panels: scatter plots with bar graphs showing the percentage of MYOD-positive and MYOG-positive nuclei on the right. The images were quantified by ImageJ v2.9 using DAPI to delimit nuclei. The cut-off point, to consider a nucleus positive for MYOD or MYOG, was set as the mean +1*SD of CTRL values. n=5-7 images per cell line. Scale bars: 50 µm. *$p<0.05$ compared to CTRL_3 via Mann-Whitney *U* test. **c** Proliferation curve measured as the fold-change in the number of DAPI-positive nuclei at days 1, 3, 5 and 7 after cell seeding at a confluence of 30%. Right panel shows the quantification at day 7 of each independent individual's cell. The inner plot depicts the comparison of the average values of each ALS patient group with the CTRL group. n=3-5 images per cell line. Bar graphs represent mean ± SEM. *$p<0.05$ compared to the CTRL group via Mann-Whitney *U* test. **d** Quantification of KI67 % over total DAPI along proliferation at days 1, 3, 5 and 7 as in **c**. Right panel shows the quantification at day 7 of each independent individual's cell. The inner plot depicts the comparison of the average values of each ALS patient group (fALS and sALS) with the CTRL group. n=3-5 images per cell line. Bar graphs represent mean ± SEM. n.s., not significant. **e** Representative Western blot images of Atrogin-1 and MURF proteins in CTRL and ALS proliferative myoblasts, and scatter dot-plots with bar graphs showing quantification of protein levels. n=3 per group. Bar graphs represent mean ± SEM. *$p<0.05$ compared to the CTRL group via Mann-Whitney *U* test.

a

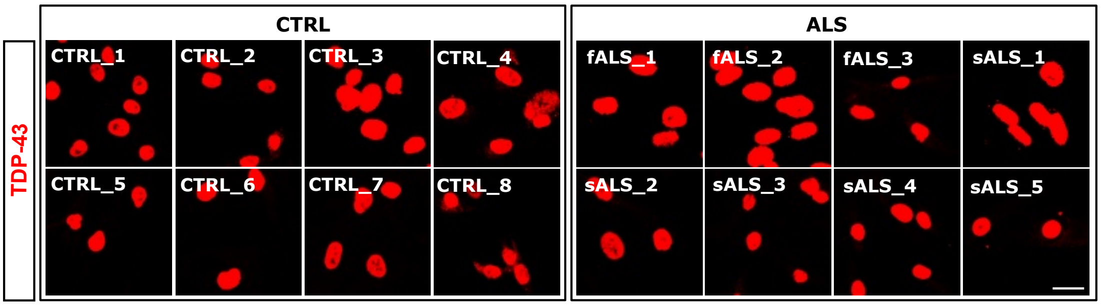


b

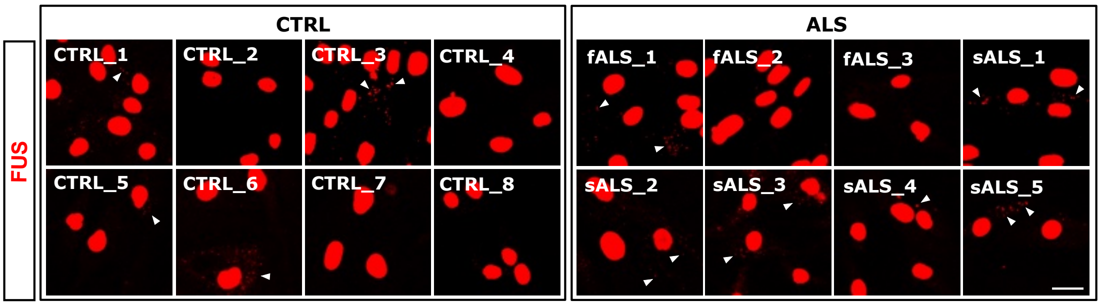


**Supplementary Figure 3. Examination of cytoplasmic granules of TDP-43 and FUS in ALS primary human myoblasts. a-b** Representative images of primary myoblasts from ALS patients and healthy controls stained for TDP-43 (**a**) or FUS (**b**). Note that the exposure levels have been saturated to enhance the visibility of the cytoplasmic granules, if present (white arrowheads). Scale bars: 25 µm. fALS, familial ALS; sALS, sporadic ALS.

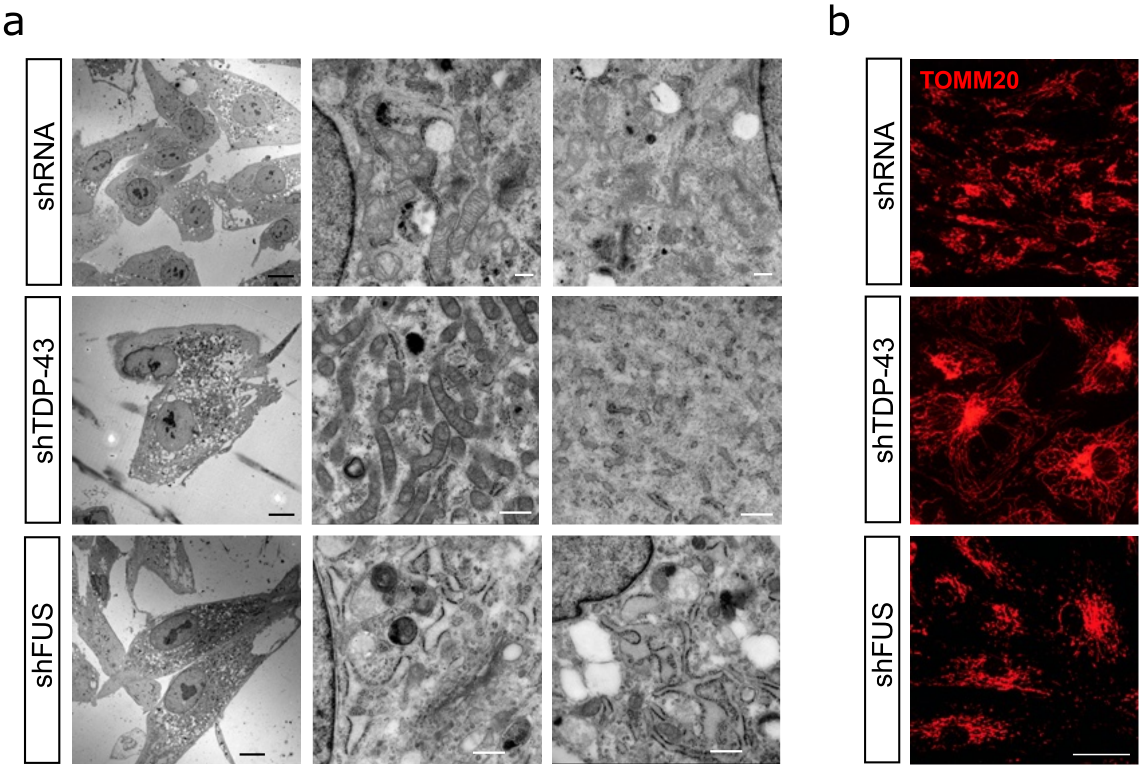


| | Mitochondria | RER | Golgi | Lysosomes | Cytoskeleton |
|---|---|---|---|---|---|
| **shRNA** | + Medium/big | + Non-dilated | +++ Short dictyosomes | ++ | ++ |
| **shTDP-43** | +++ Small | ++ Short, periphery | + | ++ | ++ |
| **shFUS** | ++ | +++ Dilated | ++ | + | ++ |

**Supplementary Figure 4. Ultrastructural characteristics of human myoblasts with TDP-43 or FUS silencing**. **a** Representative transmission electron microscopy (TEM) images of human myoblasts transduced with shRNA lentiviral particles targeting TDP-43 or FUS proteins, compared to a control group with shRNA alone. The bottom panel shows a semi-quantitative analysis of the main ultrastructural features observed, including mitochondria, rough endoplasmic reticulum (RER), Golgi apparatus, lysosomes, and cytoskeleton, across the different conditions. The symbols indicate relative abundance: "+" denotes moderate number, "++" denotes increased number, and "+++" denotes very increased number. Black scale bars represent 10 µm, and white scale bars represent 500 nm. **b** Representative immunofluorescence images of myoblasts stained with TOMM20, a mitochondrial marker, to visualize and confirm the presence of increased mitochondrial abundance in sh-TDP43 myoblasts. Scale bars: 25 µm.

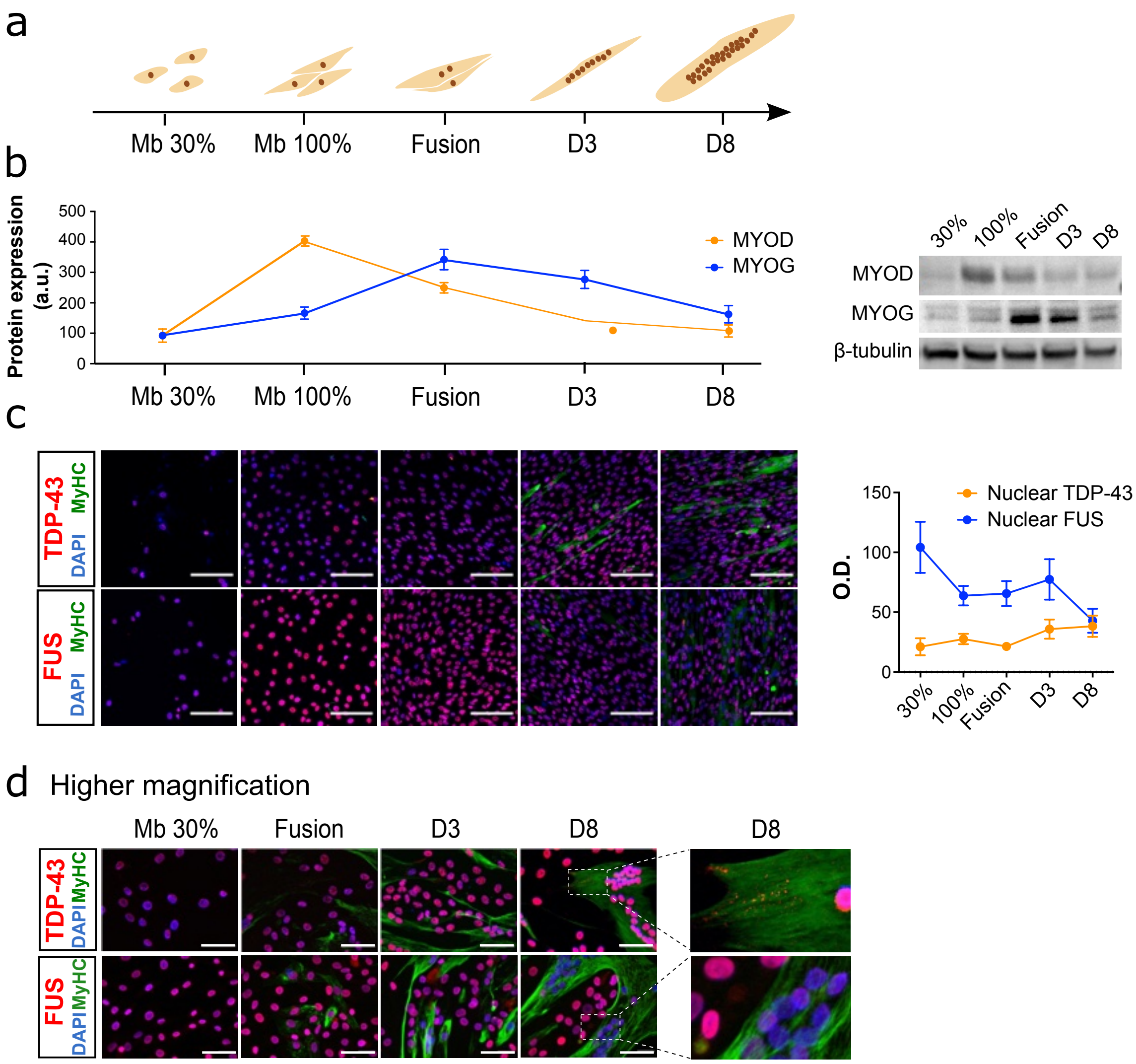


**Supplementary Figure 5. Dynamics of TDP-43 and FUS along the myogenic process in the immortalised human myoblasts. a** Schematic diagram of the myogenic process showing the different phases under study. **b** Quantification of the myogenic regulatory markers MYOD and myogenin (MYOG) protein levels throughout the myogenic differentiation process, and representative Western blot images of both proteins. Data are expressed as mean ± SEM. n=3. **c** Representative images of human immortalised myoblasts stained for TDP-43 or FUS (red) and MyHC (Myosin heavy chain, green) throughout the myogenic differentiation process, and quantification of optical densities (O.D.) of either nuclear TDP-43 or nuclear FUS staining. Nuclei were counterstained with DAPI (blue). Scale bars: 150 µm. Data are expressed as mean ± SEM. **d** Higher magnification of images of human immortalised myoblasts stained for TDP-43 or FUS (red) and MyHC (Myosin heavy chain, green) throughout the myogenic differentiation process. Scale bars: 75 µm.

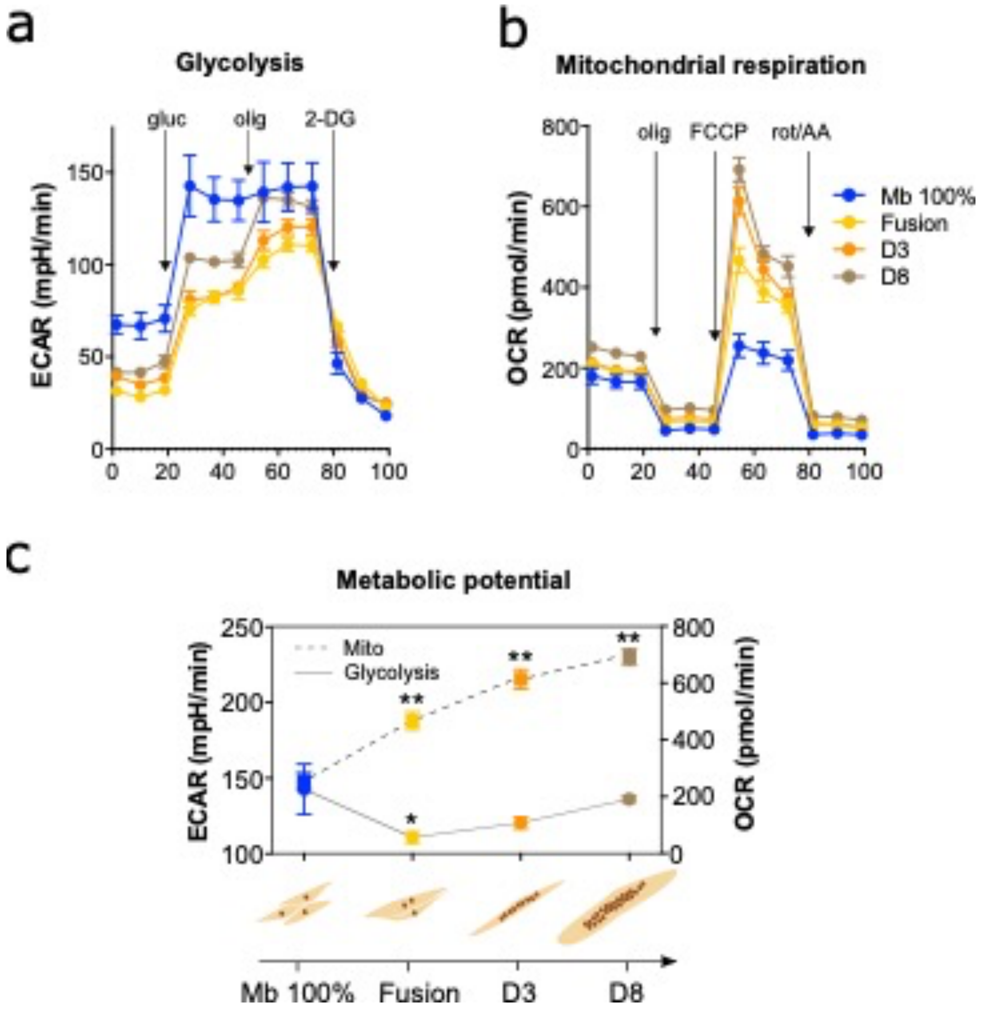


**Supplementary Figure 6. Metabolic characterization of the immortalised human myoblasts.** **a-b** Graphs showing Extracellular Acidification Rate (ECAR) (**a**) and Oxygen Consumption Rate (OCR) (**b**) of human myoblasts at different stages during the myogenic process. gluc, Glucose; olig, Oligomycin; 2-DG, 2-deoxyglucose; FCCP, Carbonyl cyanide p-(trifluoromethoxy) phenylhydrazone; rot/AA, Rotenone/Antimycin A. **c** Representation of the glycolytic capacity (Glycolysis, solid line) and the maximal respiration (Mito, dashed line) calculated by the data obtained in **a** and **b**, respectively. n=6-8 wells per group. Data are expressed as mean ± SEM. *$p$<0.05; **$p$<0.01 compared to the myoblasts at 100% confluence (Mb 100%) via one-way ANOVA.

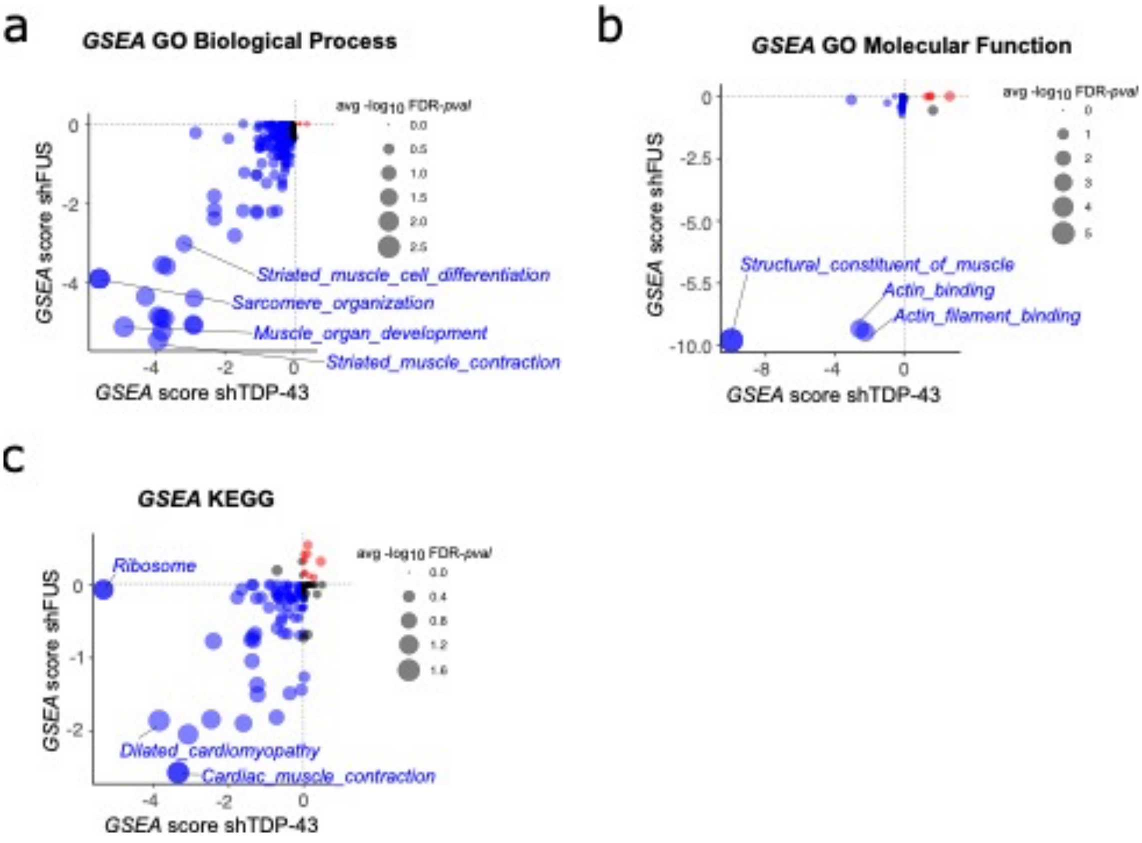


**Supplementary Figure 7. Extended Figure 4c. a-c** Bubble charts of ranked enrichment analysis of shTDP-43 plotted against shFUS gene expression signatures using the GO Biological Process (**a**), GO Molecular Function (**b**) and KEGG (**c**) databases by the GSEA multilevel enrichment test. GSEA score rank is based on normalised enrichment score (NES) and −log10FDR-pval for each gene set. Bubble size represents the average −log10FDR *p*-value. Gene sets upregulated and downregulated in both shTDP-43 and shFUS myoblasts are indicated in red and blue, respectively.

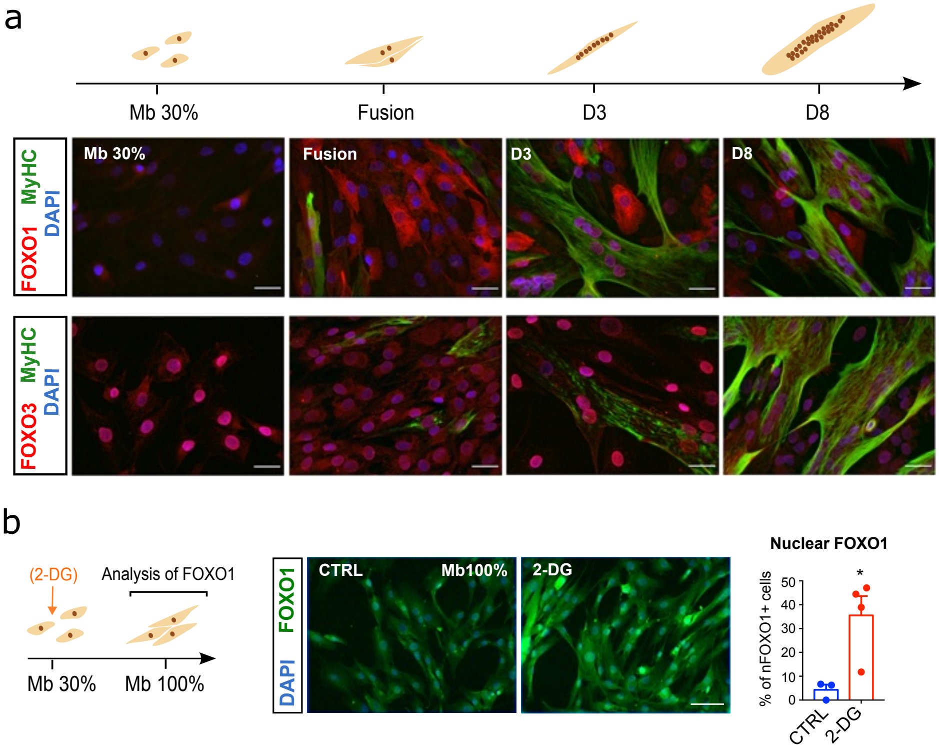


**Supplementary Figure 8. Dynamics of FOXO transcription factors throughout myogenesis and metabolic challenges in human myoblasts. a** Representative images of human immortalised myoblasts stained for FOXO1 (red) or FOXO3 (green) and MyHC (Myosin heavy chain, green or red) and DAPI (nuclei, blue) throughout the myogenic differentiation process. Scale bars: 25 µm. **b** Representative images of 2-deoxyglucose (2-DG)-treated myoblasts at 100% confluence (Mb 100%) stained for FOXO1 (green) and DAPI (blue), and scatter dot-plot with a bar graph showing the percentage of cells positive for nuclear FOXO1 staining, which were quantified by ImageJ v2.9 software using DAPI staining to delimit nuclei. Nuclei with integrated density values above the cut-off point (set as the value of mean +1*SD of CTRL cells) were considered positive for FOXO1. Scale bar: 50 µm. n=3-4 images per group. *$p$<0.05 compared to the untreated control (CTRL) group via Mann-Whitney $U$ test.

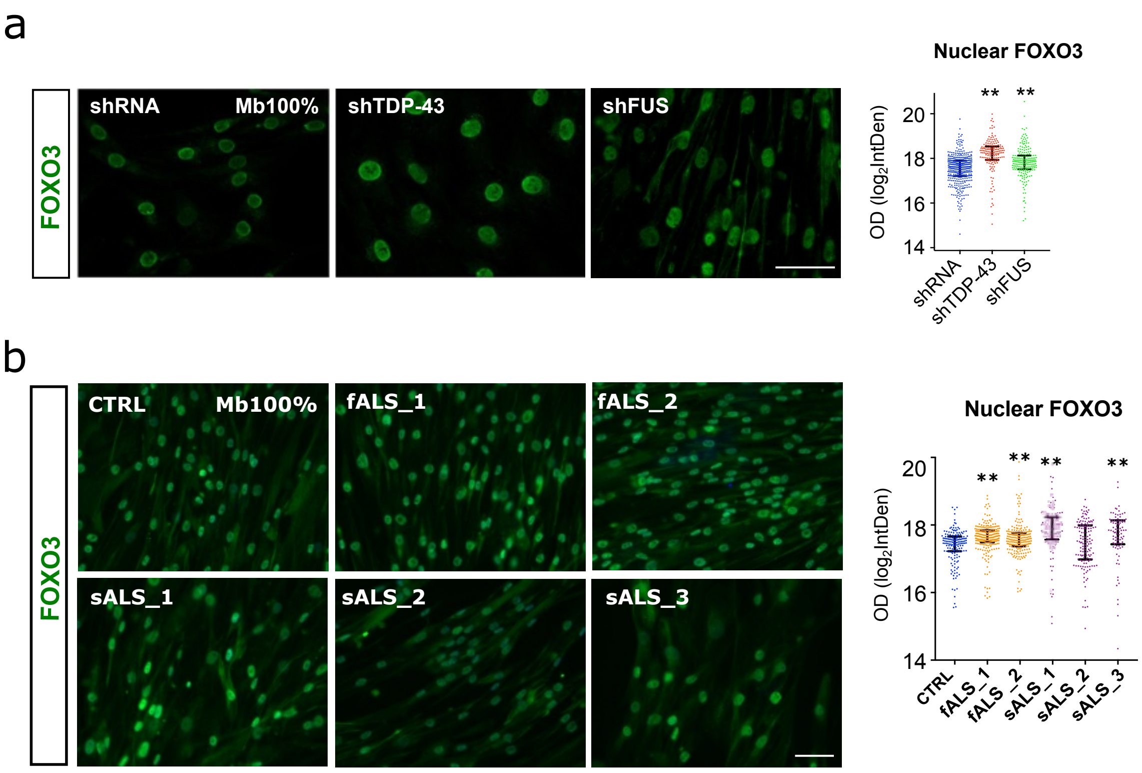


**Supplementary Figure 9. Levels of nuclear FOXO3 in myoblasts models of ALS. a** Representative images of TDP-43 or FUS-knockdown myoblasts at 100% confluence (Mb100%) stained for FOXO3 (green). On the right, quantitative scatter dot-plots with median and interquartile range for the integrated densities of nuclear FOXO3 staining, which were quantified by ImageJ v2.9 software using DAPI staining to delimit nuclei. Scale bar: 50 µm. n=3 images per group. ***$p<0.01$ compared to the shRNA control group via one-way ANOVA. **b** Representative images of a selection of ALS myoblasts and an age-matched CTRL myoblast at Mb100% stained for FOXO3 (green). On the right, quantitative scatter dot-plots with median and interquartile range for the integrated densities of nuclear FOXO3 staining, which were quantified by ImageJ v2.9 software using DAPI staining to delimit nuclei. Scale bar: 100 µm. n=3 images per group. ***$p<0.01$ compared to the CTRL group via one-way ANOVA. Scale bar: 50 µm. fALS, familial ALS; sALS, sporadic ALS. OD, optical densities.

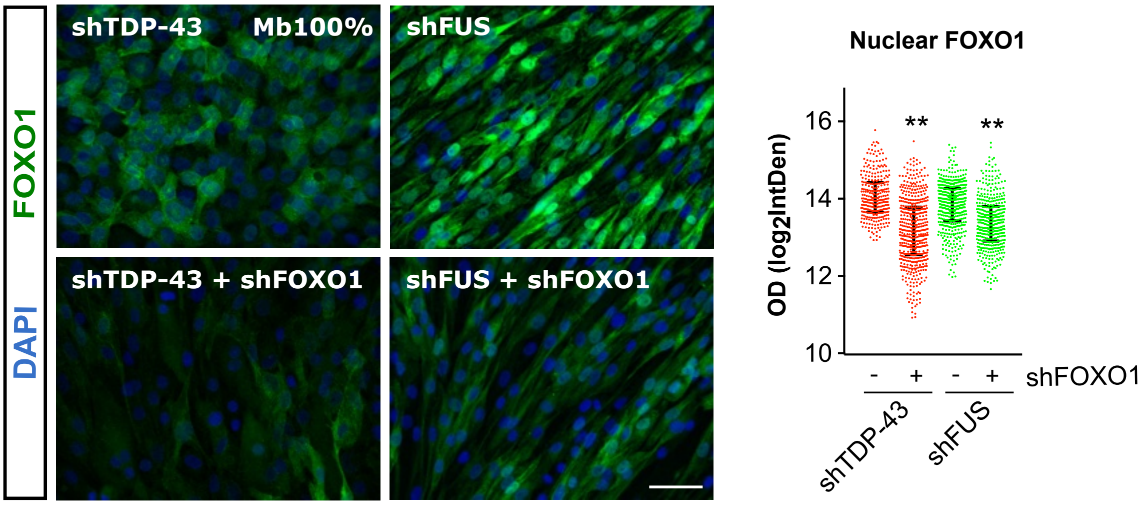


**Supplementary Figure 10. Effect of FOXO1 silencing on the levels of nuclear FOXO1 in human myoblasts silenced for TDP-43 or FUS.** Representative images of TDP-43 or FUS-knockdown myoblasts at 100% confluence (Mb100%) treated or not with shFOXO1 lentiviral particles stained for FOXO1 (green). On the right, quantitative scatter dot-plots with median and interquartile range for the integrated densities of nuclear FOXO1 staining, which were quantified by ImageJ v2.9 software using DAPI staining to delimit nuclei. Scale bar: 50 µm. n=3 images per group. ***p*<0.01 compared to cells not treated with shFOXO1 particles via one-way ANOVA. OD, optical densities.

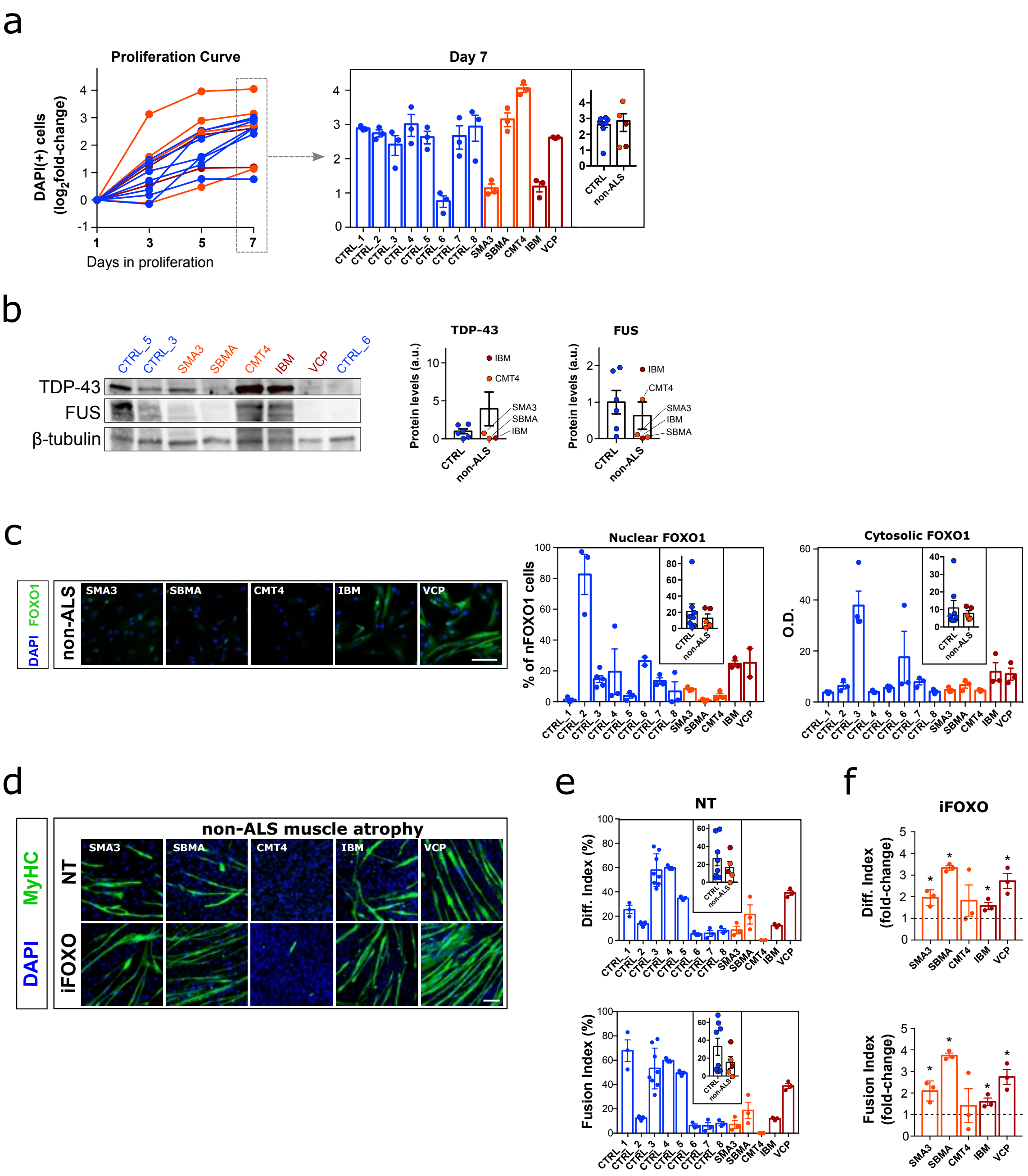


**Supplementary Figure 11. FOXO1 is not dysregulated in primary myoblasts from non-ALS neurogenic atrophies and myopathies. a** Proliferation curve measured as the fold-change in the number of DAPI-positive nuclei at days 1, 3, 5, and 7 after cell seeding at a confluence of 30%. Right panel shows the quantification at day 7 of each independent individual's cell. The inner plot depicts the comparison of the average values of each non-ALS neurogenic atrophy (SMA3, SBMA, CMT4) or myopathy (IBM, VCP-related) with the healthy control group (CTRL). n=3-5 images per cell line. Bar graphs represent mean ± SEM. **b** Representative Western blot images of TDP-43 and FUS proteins in myoblasts from CTRL and non-ALS patients, and scatter dot-plots with bar graphs showing the quantification of protein levels. n=3 per group. **c** Left panel: representative images of primary myoblasts at 100% confluence from patients with non-ALS neurogenic atrophy or myopathy and healthy CTRL stained for FOXO1 (green) and DAPI (nuclei, blue). Right panel shows a dual representation of FOXO1 staining: scatter plot showing the distribution of cells positive for nuclear FOXO1, and bar graph depicting the mean optical density (OD) of cytosolic FOXO1 staining. Quantifications were made by ImageJ v2.9 software using DAPI staining to delimit nuclei. Nuclei with integrated density values above the cut-off point (set as the value of mean +1*SD of CTRL cells) were considered positive for FOXO1. The inner plot depicts the comparison of the average values of each patient group (non-ALS neurogenic atrophy or myopathy) with the average values of the CTRL group. Scale bars: 50 µm. n=3-5 images per group. Bar graphs represent mean ± SEM. **d** Representative images of primary myoblasts from patients with non-ALS neurogenic atrophy or myopathy and CTRL treated or not (NT) with the selective FOXO1 inhibitor AS1842856 (iFOXO1) at 30 nM and stained for MyHC (Myosin heavy chain, green) and DAPI (nuclei, blue). Immunofluorescence was performed 8 days (D8) after the first fusion events of NT CTRL cells. Scale bars: 100 µm. n=5-6 images per group. **e** Quantitative scatter plots with bar graphs showing fusion and differentiation indices from n=3 independent differentiation assays. The inner plot depicts the comparison of the average values of each patient group (non-ALS neurogenic atrophy or myopathy) with the average values of the CTRL group. **f** Quantitative scatter plot with bar graphs showing the fold-change of treated cells relative to the untreated ones (dotted line). Bar graphs represent mean ± SEM. SMA3, Spinal Muscular Atrophy type 3; SBMA, Spinal Bulbar Muscular Atrophy; CMT4, Charcot-Marie-Tooth type 4; IBM, sporadic Inclusion Body Myositis; VCP, vacuolar myopathy related to Valosin-Containing Protein mutation.

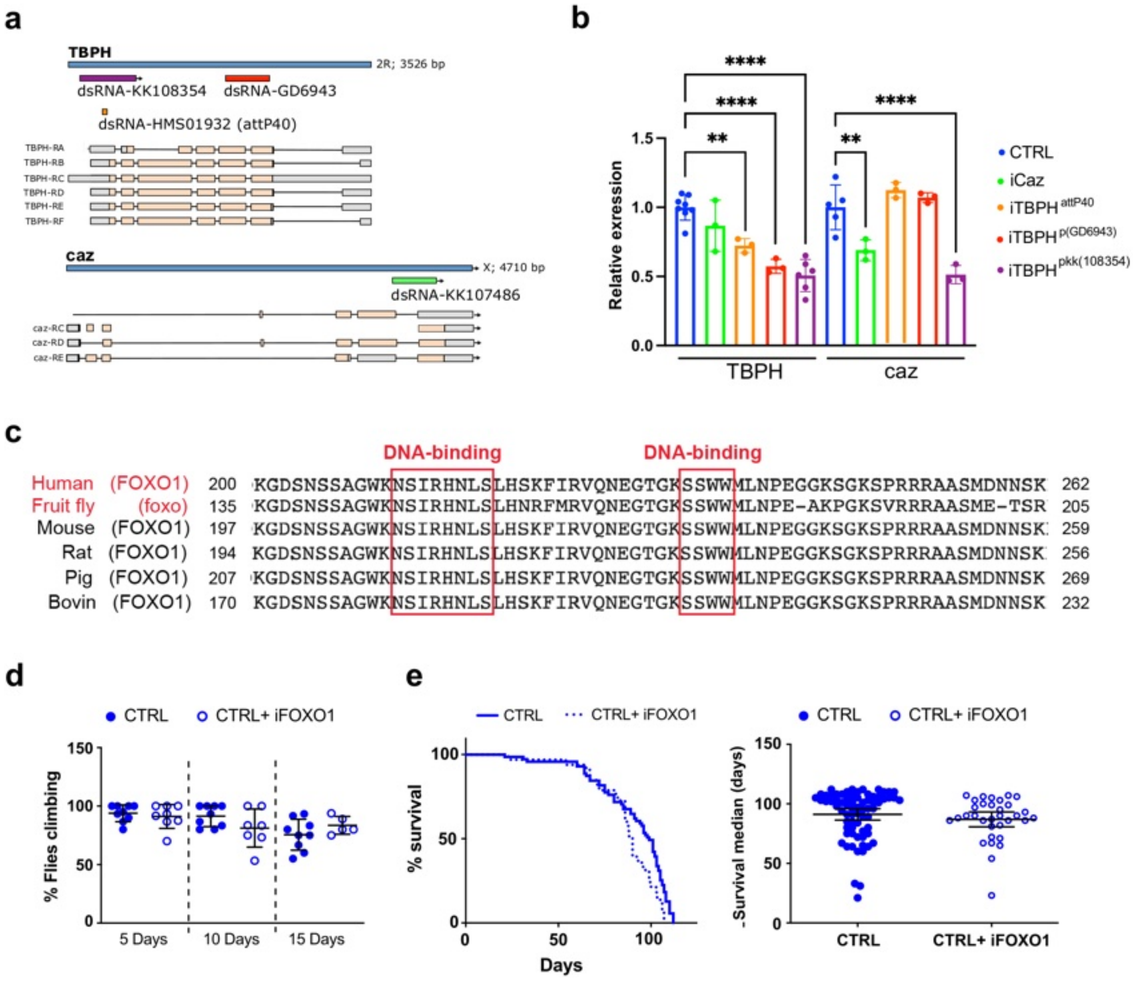


**Supplementary Figure 12. Extended Fig. 7. a** Gene schematic diagrams illustrating the size and location of sequences used in each fly model to generate dsRNA for RNA interference of *TBPH* or *caz* genes (human *TARDBP* or *FUS* orthologs, respectively). **b** Quantitative scatter plots with bar graphs showing expression of *TBPH* and *caz* genes measured by quantitative PCR in ALS *Drosophila* models (with muscle-specific silencing) relative to control (CTRL) flies. Bar graphs represent mean ± SEM. n=3-5 flies. **$p$<0.01; ****$p$<0.0001 compared to CTRL flies via one-way ANOVA. **c** Multiple sequence alignment for FOXO1 across species, highlighting the conserved DNA binding sites that are targeted by the AS1842856 FOXO1 inhibitor (iFOXO). **d** Scatter dot-plots with median and interquartile range showing climbing activity of CTRL flies treated or not with iFOXO1 on adult days 5, 10 and 15. n=5 flies per tube, 5-9 tubes per group. Each value is calculated as the average of three trials. **e** Kaplan-Meier curves of CTRL flies treated or not with iFOXO1 (left panel) and scatter dot-plots with median and interquartile range showing median survival times (right panel). n=100 flies per group; **$p$<0.01 compared to the untreated flies via log-rank test.
